# Improved Robustness of SARS-CoV-2 Whole-Genome Sequencing from Wastewater with a Nonselective Virus Concentration Method

**DOI:** 10.1101/2022.09.07.22279692

**Authors:** Emily Segelhurst, Jonathan E. Bard, Annemarie N. Pillsbury, Md Mahbubul Alam, Natalie A. Lamb, Chonglin Zhu, Alyssa Pohlman, Amanda Boccolucci, Jamaal Emerson, Brandon J. Marzullo, Donald A. Yergeau, Norma J. Nowak, Ian M. Bradley, Jennifer A. Surtees, Yinyin Ye

## Abstract

The sequencing of human virus genomes from wastewater samples is an efficient method for tracking viral transmission and evolution at the community level. However, this requires the recovery of viral nucleic acids of high quality. We developed a reusable tangential-flow filtration system to concentrate and purify viruses from wastewater for whole-genome sequencing. A pilot study was conducted with 94 wastewater samples from four local sewersheds, from which viral nucleic acids were extracted, and the whole genome of severe acute respiratory syndrome coronavirus 2 (SARS-CoV-2) was sequenced using the ARTIC V4.0 primers. Our method yielded a high probability (0.9) of recovering complete or near-complete SARS-CoV-2 genomes (>90% coverage at 10× depth) from wastewater when the COVID-19 incidence rate exceeded 33 cases per 100 000 people. The relative abundances of sequenced SARS-CoV-2 variants followed the trends observed from patient-derived samples. We also identified SARS-CoV-2 lineages in wastewater that were underrepresented or not present in the clinical whole-genome sequencing data. The developed tangential-flow filtration system can be easily adopted for the sequencing of other viruses in wastewater, particularly those at low concentrations.

**SYNOPSIS:** The tangential-flow filtration method extracts viral nucleic acids of high enough quality from wastewater for robust and successful whole-genome sequencing.

**GRAPHIC FOR TABLE OF CONTENTS (TOC):** 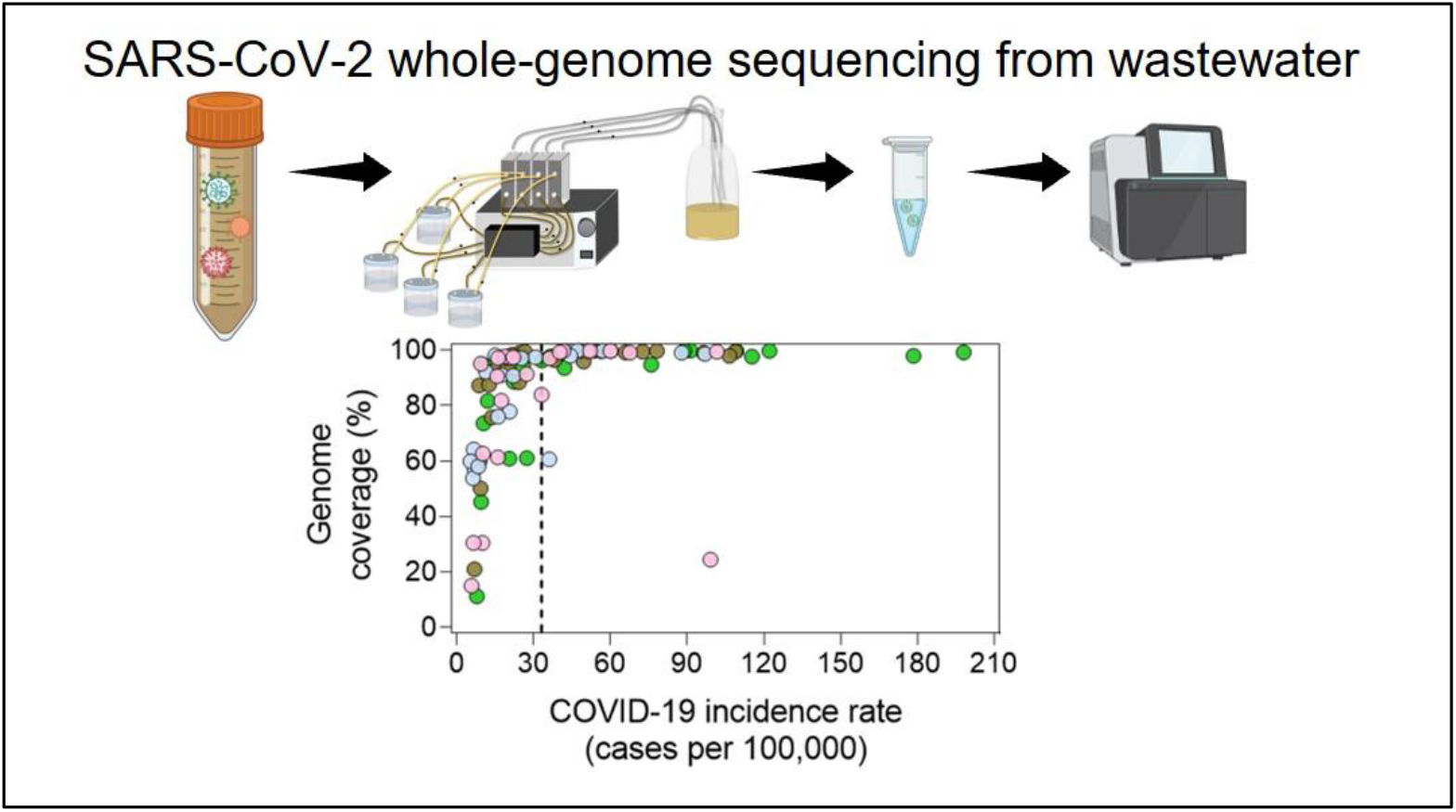

## INTRODUCTION

The global spread of severe acute respiratory syndrome coronavirus 2 (SARS-CoV-2) has facilitated the emergence of genome mutations, resulting in new lineages that further threaten public health.^1^ Whole-genome sequencing (WGS) of clinical samples is a powerful method for tracking the spread of various SARS-CoV-2 lineages,^2^ but this can be expensive, slow, and subject to biased sampling. Furthermore, the frequency of clinical testing has declined, aided by the availability of rapid at-home testing kits. WGS of SARS-CoV-2 in wastewater samples circumvents the issues as a high-throughput and more comprehensive strategy that monitors a larger portion of the population.^3-6^ Successful WGS of SARS-CoV-2 in wastewater samples relies on the depth (the number of times a nucleotide is sequenced)^7^ and breadth of genome coverage (hereafter, coverage; the percentage of nucleotide positions sequenced to a given depth).^7^ Because SARS-CoV-2 lineages differ by only a few mutations, greater depth and coverage are needed to quantify lineage abundance and detect low-frequency mutations.^8, 9^

Different virus concentration methods have been developed for polymerase chain reaction (PCR)-based detection of viral genes from wastewater.^10-13^ However, the nucleic acids recovered with these methods may not be suitable for WGS—PCR assays typically target <1% of the genome. Sequencing inhibitors, such as humic acids, and background nucleic acids from prokaryotes and eukaryotes are likely to reduce the genome coverage of SARS-CoV-2. Indeed, near-complete (>90%) SARS-CoV-2 genomes are not recovered from most samples (>85%) when aluminum hydroxide is used to directly precipitate viruses without separating viruses from other microorganisms in wastewater.^14^ The removal of wastewater solids before the Amicon/Centricon ultra-centrifugal concentration^15^ or electronegative membrane filtration^16^ can improve sequencing success, but the quality of the extracted genomes can vary and random sequencing failures can occur.^17^

We report a reusable and nonselective tangential-flow filtration system to concentrate and purify viruses from wastewater for WGS. We evaluated the robustness of the method to recover complete or near-complete SARS-CoV-2 genomes using the samples collected from four local sewersheds over six months. We subsequently compared the findings with those from patient-derived clinical samples at the County level. Finally, the timing of lineage detection was compared among clinical WGS, wastewater WGS, and quantitative reverse transcription polymerase chain reactions (RT-qPCR).

## MATERIALS AND METHODS

### Wastewater sample collection

Time- or flow-weighted 24-hr composite influent samples were collected every 1 or 2 weeks between October 1 and December 16, 2021, and between January 18 and April 12, 2022, from three wastewater treatment plants covering four sewersheds that serve ∼80% of the total population of Erie County (New York): Tonawanda, Kenmore-Tonawanda, Amherst, and Bird Island (**Table S1**; **Figure S1**). Influent was collected by an autosampler every 30 min, and the samples were kept at 4 °C in high-density polyethylene bottles that were cleaned with 10% bleach. The samples were transported on ice packs and stored at 4 °C until they were processed.

### Virus concentration from wastewater

Wastewater samples (125 mL) were centrifuged in sterile bottles at 10 000 × *g* for 15 min at 4 °C to remove large wastewater solids, prokaryotic and eukaryotic cells, and other debris. The supernatant was concentrated in a Vivaflow laboratory cross flow cassette system (Sartorius) equipped with a 30-kDa Hydrosart ultrafilter membrane (Sartorius) at a feedline flow rate of 8.5 mL/min (**Figure S2**). The residual liquid in the ultrafilter membrane and connected tubing was blown with air into the sample reservoir. The concentrate (∼25 mL) in the reservoir was then collected, overlayed with 5 mL of a 20% (v/v) sucrose solution and ultracentrifuged at 100 000 × *g* for 45 min at 4 °C in an SW 32 Ti rotor using an Optima XE series centrifuge (Beckman Coulter). Immediately after the ultracentrifugation, the pellet was resuspended in 200 μL phosphate-buffered saline (pH 7.4; Gibco) and stored at −80 °C ∼two weeks until nucleic acid extraction.

After use, the ultrafilter membranes were immediately flushed with ∼100 mL of 0.5 M NaOH solution preheated to 55 °C at a feedline flow rate of 8.5 mL/min. The NaOH solution was then recirculated for 20–30 min at the same flow rate. The cleaned membrane was stored in 0.5 M NaOH at 4 °C. Cleaning efficiency was verified on six random days by processing 125 mL of autoclaved Milli-Q water with the same concentration procedure. The absence of SARS-CoV-2 genes through washed filters were confirmed by RT-qPCR (**Figure S3**).

### Nucleic acid extraction and RT-qPCR

Viral nucleic acids were extracted from the concentrated samples with QIAamp viral RNA mini kits (Qiagen) and eluted in 60 µL AVE buffer (Qiagen) per the manufacturer’s instructions. One-step RT-qPCR was performed to quantify the SARS-CoV-2 N gene according to the CDC N2 assay^18^ and to determine the presence/absence of specific S gene mutations, including WT493-498, Q493R and Q498R, delH69/V70, and delL24/P25/P26 and A27S as variant determinants for Delta, generic Omicron, Omicron BA.1, and Omicron BA.2, respectively, following methods published previously (**Table S2**).^19, 20^ The limit of detection for the SARS-CoV-2 N gene was determined to be 1.8 gene copies/µL, and the limit of quantification was determined to be 5 gene copies/µL following a method as described previously (**Figure S4**).^21^ A checklist of the MIQE (<u>m</u>inimum information for publication of <u>q</u>uantitative real-time PCR <u>e</u>xperiments) guidelines^22^ is provided in **Table S3**. All RT-qPCR assays were conducted in duplicates on a CFX96 Touch real-time PCR detection system (Bio-Rad), and the threshold cycle (*C*^*T*^) value was determined using a CFX Maestro Software (Version 4.0, Bio-Rad). To remove PCR inhibition, the extracted nucleic acids were diluted 5- or 10-fold in nuclease-free water before RT-qPCR analyses (**Figure S5**). Detailed procedures of RT-qPCR inhibition test, reaction compositions, thermocycling conditions, and determination of the limits of detection and quantification are described in the Supporting Information.

### SARS-CoV-2 whole-genome sequencing (WGS)

The quality of the extracted nucleic acids was assessed with the Agilent Fragment Analyzer. WGS of SARS-CoV-2 was performed following a modified ARTIC protocol^23^ using the V4.0 nCOV-2019 amplicon panel (IDT). Briefly, 8 µL of nucleic acid extracts was reverse transcribed to cDNA with random hexamers using the Invitrogen SuperScript IV first-strand synthesis system (Thermo Fisher Scientific) according to the manufacturer’s protocol. The cDNA was amplified with two primer pools using Q5 hot-start high-fidelity 2× master mix (New England BioLabs) with the following parameters: 98 °C for 30 s, 25 cycles of 98 °C for 15 s and 65 °C for 5 min. The resulting amplicons from both primer pools were combined. Excess primers and reagents were removed with 1× AMPure XP beads (Beckman Coulter), and the amplicons were eluted in EB buffer (Qiagen). The total eluted volume of amplicons was input to generate libraries using the NEBNext Ultra II DNA library prep kit (New England BioLabs) without fragmentation per the manufacturer’s protocol. Individual samples were then barcoded and pooled for quantification using the sparQ Universal Library Quant kit (QuantaBio) for Illumina sequencing. The libraries were diluted to 10 pM and sequenced on an Illumina MiSeq instrument (V3 chemistry, PE300) with 1% PhiX as internal control.

### Bioinformatic analysis

The Illumina sequencing output files were demultiplexed using bcl2fastq (v2.20.0.422) to convert into FASTQ files. Initial quality control was performed through FastQ Screen.^24^ Samples were then processed using the UB Genomics and Bioinformatics Core SARS-CoV-2 analysis pipeline (https://github.com/UBGBC/fastq-to-consensus). Briefly, adapters were trimmed before sequencing reads were aligned to the SARS-CoV-2 reference genome (MN908947.2) using the BWA-MEM algorithm.^25^ Variants, insertions, and deletions were then called using BCFtools (v.1.10.2),^26^ requiring a minimum depth per nucleotide position of 10× or 50× and generating VCF file outputs along with a final consensus FASTA file for each input sample. The resulting VCF and depth mpileup files were used as input into Freyja to perform lineage composition analysis (https://github.com/andersen-lab/Freyja).5 The coverage (--covcut) was specified at 10× or 50×, and only confirmed lineages were reported. The Freyja pipeline was selected because of its high accuracy and efficiency.^5^

### Patient-derived SARS-CoV-2 data

The daily cases of COVID-19 during the sampling periods in the studied sewersheds were extracted from the Erie County Wastewater Monitoring Dashboard.^27^ The COVID-19 incidence rates (per 100 000 population in the sewershed) were then calculated as 7-day rolling averages. The patient-derived WGS data at the County level were downloaded from the Global Initiative on Sharing All Influenza Data (GISAID) database^28^ with the location as “North America/USA/New York/Erie County,” collection date from “2021-10-01” to “2022-04-30”, and low coverage excluded (GISAID Identifier: EPI_SET_220906wr). Most of these samples were sequenced at University at Buffalo.^29^ On each collection date, the relative abundances of SARS-CoV-2 lineages in the patient samples were calculated, and the number of patient samples that were collected for sequencing were smoothed as 7-day rolling averages.

### Statistical analysis

The probability of recovering >90% coverage of the SARS-CoV-2 genome at 10× depth at a given RT-qPCR *C*_*T*_ value or COVID-19 incidence rate was predicted by binary logistic regression analysis. Data were visualized with Prism 9.2.0 (GraphPad Software) and the ggplot2^30^ package in R version 4.2.1.^31^

### Data availability

Raw sequencing data are available in NCBI Sequence Read Archive (SRA) under the BioProject ID: PRJNA877272. Codes for analyzing SARS-CoV-2 lineage in wastewater are available at https://github.com/UBGBC/fastq-to-consensus.

## RESULTS AND DISCUSSION

### Coverage and depth of SARS-CoV-2 genomes from wastewater

Complete or near-complete SARS-CoV-2 genomes (>90% coverage) at 10× depth were recovered from 68% (64/94) of the wastewater samples (**Table S4**). This success rate is 53% higher than that in a SARS-CoV-2 wastewater sequencing study that applied aluminum hydroxide flocculation and the ARTIC protocol.^14^ A binary logistic regression analysis predicted a 0.9 probability of sequencing success (>90% coverage at 10× depth) when the *C*_*T*_ value of the SARS-CoV-2 N gene was 31.8 (corresponding to ∼3500 gene copies loaded for sequencing) (**Figure 1A**). For comparison, at the 0.9 probability of sequencing success, *C*_*T*_ values of SARS-CoV-2 gene ranged from <26 to ∼29 with other workflows in previous pilot studies (**Figure S6**).^14-17^ Our workflow demonstrates higher coverage of SARS-CoV-2 genome at higher *C*_*T*_ values (lower gene levels).

**Figure 1.**
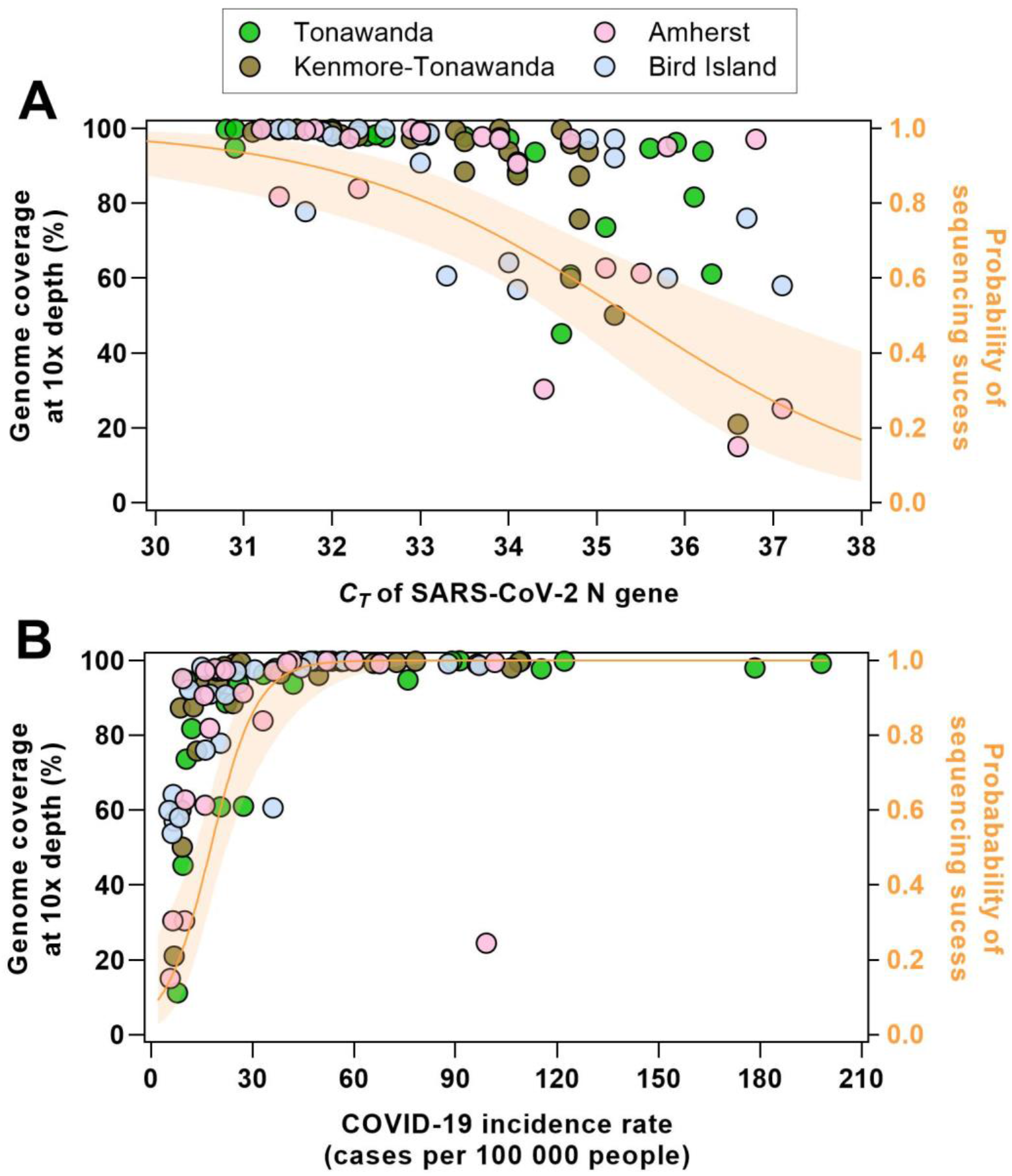
Coverage of the SARS-CoV-2 genome at 10× depth and the probability of sequencing success (>90% coverage at 10× depth) with regard to *C*_*T*_ value of the SARS-CoV-2 N gene measured with RT-qPCR (A) and the COVID-19 incidence rate (rolling 7-day average of cases per 100 000 people) (B). The probability of sequencing success was calculated according to a binary logistic regression analysis.

Most genomic regions were sequenced at a minimum depth of 50× (**Figure S7**). We noted that three regions were often sequenced at lower depth (<10×): nucleotides 21750 to 22250 in the S gene (amplicons 72 and 73) and 26750 to 27000 in the M gene (amplicon 89) (**Figure S7**). Similar amplicon dropout issues were previously reported for a few amplicons using the ARTIC V3 panel.^32, 33^ We used ARTIC V4.0 primers, which address the V3 coverage issues but do not include all the mutations in SARS-CoV-2 Delta and Omicron lineages in the primer design.^34^ Other wastewater factors may also explain the low-depth coverage within certain regions of SARS-CoV-2 genome. First, the low concentrations of the SARS-CoV-2 genome in wastewater (<10^5^ gene copies/mL)^35^ may have contributed to the lower sequencing depth. The presence of primer dimers compete for primer-template interactions, resulting in reduced amplification efficiency.^36^ Also, mutations of SARS-CoV-2 genomes identified in wastewater but not in patient samples could influence primer binding at some sites.^3, 6, 37^ Our results suggest that up-to-date primer design and optimization is critical to sequence emerging variants from wastewater.

### Robustness of SARS-CoV-2 WGS from wastewater

#### Influence of sewersheds

The rates of sequencing success were similar for samples collected from the four studied sewersheds: 70.8% (17/24) for Tonawanda, 72.0% (18/25) for Kenmore-Tonawanda, 72.7% (16/22) for Amherst, and 65.2% (15/23) for Bird Island (**Table S4**). Although the Bird Island sewershed serves a population that is larger than the other three (**Table S1**), similar average *C*_*T*_ values were obtained for the SARS-CoV-2 N gene (33.5 for Tonawanda, 33.4 for Kenmore-Tonawanda, 34.2 for Amherst, and 34.1 for Bird Island) (**Table S4**), indicating that similar genome levels were available for sequencing. Notably, the Bird Island wastewater treatment plant collects wastewater exclusively through combined sewer systems (**Table S1**). A large fraction of sequencing inhibitors, such as humic acids and heavy metals, in the stormwater runoff may slightly reduce the WGS success rate.^38, 39^

#### Influence of COVID-19 incidence

The probability of sequencing success was >0.9 when the COVID-19 incidence rate was >33/100 000 persons but decreased to 0.75 when the incidence rate was 25.2/100 000 persons (**Figure 1B**). A previous study reported a probability of 0.75 to quantify SARS-CoV-2 gene in wastewater by RT-qPCR when the 14-day COVID-19 case rate was 152/100 000 persons.^40^ In that study, viruses were concentrated from solids-removed influent samples by 10-kDa Centricon ultra-filters.^40^ We are not aware of other wastewater sequencing studies that have calculated SARS-CoV-2 WGS success rates with regard to the incidence rate, but our results suggest that SARS-CoV-2 WGS with the viruses recovered by tangential-flow filtration method can be successfully applied at low COVID-19 incidence rates.

### Lineage distributions estimated by wastewater and clinical data

Most of our wastewater samples were sequenced to an average depth of >50× per 250-nucleotide-region across the genome (**Figure S7**). Although the estimated lineage abundances at sequencing depths of 10× and 50× were very similar (**Figure S8**), we only reported lineage distributions estimated by the WGS data filtered at a depth of 50×.

Nine dominant groups of lineages were identified in the wastewater samples: AY.* (Delta), B.1.617.2 (Delta), B.1.2, B.1.1.529 (Omicron), BA.1/BA.1.1 (Omicron), BA.1.1.16 (Omicron), other BA.1.* (Omicron), BA.2.12/BA.2.12.1 (Omicron), and other BA.2.* (Omicron). The prevalence of Delta AY.* lineages in wastewater in October–December 2021 was coincident with the lineages observed via clinical surveillance (**Figure 2**). Two Omicron infection waves in 2022 (BA.1 in January/February and BA.2 in March/April) predicted from the wastewater data aligned with clinical data (**Figure 2**). Remarkably, wastewater sequencing revealed lineages that were underrepresented or not present in the clinical data. Specifically, the B.1.2 lineage was abundant in the Kenmore-Tonawanda sewershed in October but was not detected in patient samples from that time (**Figure 2A**). Moreover, in January/February of 2022, the BA.1/BA.1.1 lineages were prevalent in patient samples (**Figure 2A**), whereas the BA.1.1.16 lineage dominated in the wastewater samples (**Figure 2B–E**). Notably, the genome sequences of the BA.1/BA.1.1 and BA.1.1.16 lineages are similar, which might affect the predictions of their relative abundance.

**Figure 2.**
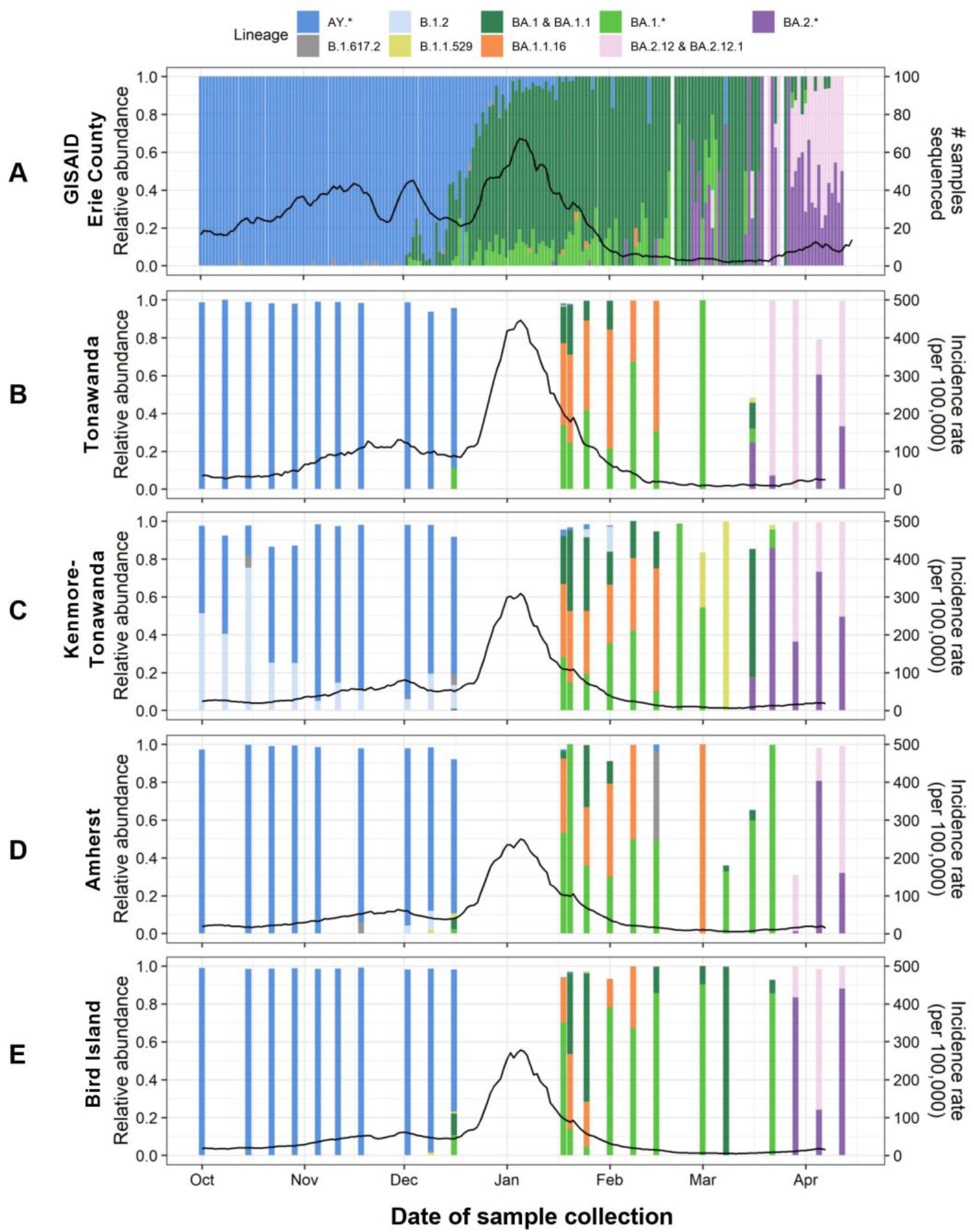
Distributions of dominant SARS-CoV-2 lineages estimated from Erie County clinical data deposited on GISAID (A) and wastewater sequencing data (B–E) during the sampling periods. The solid black lines represent the rolling 7-day average numbers of clinical samples collected for sequencing (A) and COVID-19 incidence rates (cases per 100 000 people) in each sewershed (B– E).

The reason that B.1.2 lineage identified in wastewater samples but largely absent from patient samples is unclear. The Freyja pipeline identified B.1.2 on the basis of a mutation spectrum, consisting of 13 nucleotide substitutions (A18424G, A23403G, C10319T, C1059T, C14408T, C21304T, C241T, C27964T, C28472T, C28869T, C3037T, G25563T, and G25907T), three of which (C1059T, C21304T, and G25563T) overlap the mutations in highly abundant Delta lineages circulating at the time. Furthermore, the B.1.2 mutation spectrum does not share a strong correlation with any of the mutation spectra of Delta lineages (**Figure S9**). This is less likely a misassignment by the pipeline. Taken together, our findings suggest that the WGS of patient samples can miss SARS-CoV-2 lineages circulating in the community. Those lineages may not be clinically relevant, but future research is needed to understand their roles in viral evolution and lineage emergence.

### Early detection of variants in wastewater

The values of early detection of SARS-CoV-2 variants in wastewater depend on the delays in clinical analysis.^41^ The workflow optimization for a quick turnaround of WGS analysis is beyond the scope of this study. Here, we only compare the detection of emerging SARS-CoV-2 variants based on the date of collection for both wastewater and patient samples.

The Omicron BA.1 was first identified in Amherst and Bird Island wastewater with a relative abundance of 1-2% in the second week of December, which is ∼1 week later than the patient-derived samples (**Figure 3**). The detection of Omicron BA.2 from wastewater samples in March was similarly delayed (**Figure 3**). Note that some BA.2 patient-derived samples were collected in January and February, but there was no Omicron BA.2 outbreak until March. The poor detection of Omicron BA.2 in wastewater may be attributable to the relatively low coverage of the SARS-CoV-2 genome from wastewater during mid-February and early March (<65%; Table S3). Nevertheless, our findings suggest that wastewater is a relevant proxy for patient samples. Wastewater WGS should be considered in the face of practical delays in clinical sequencing for early detection of variants.

**Figure 3.**
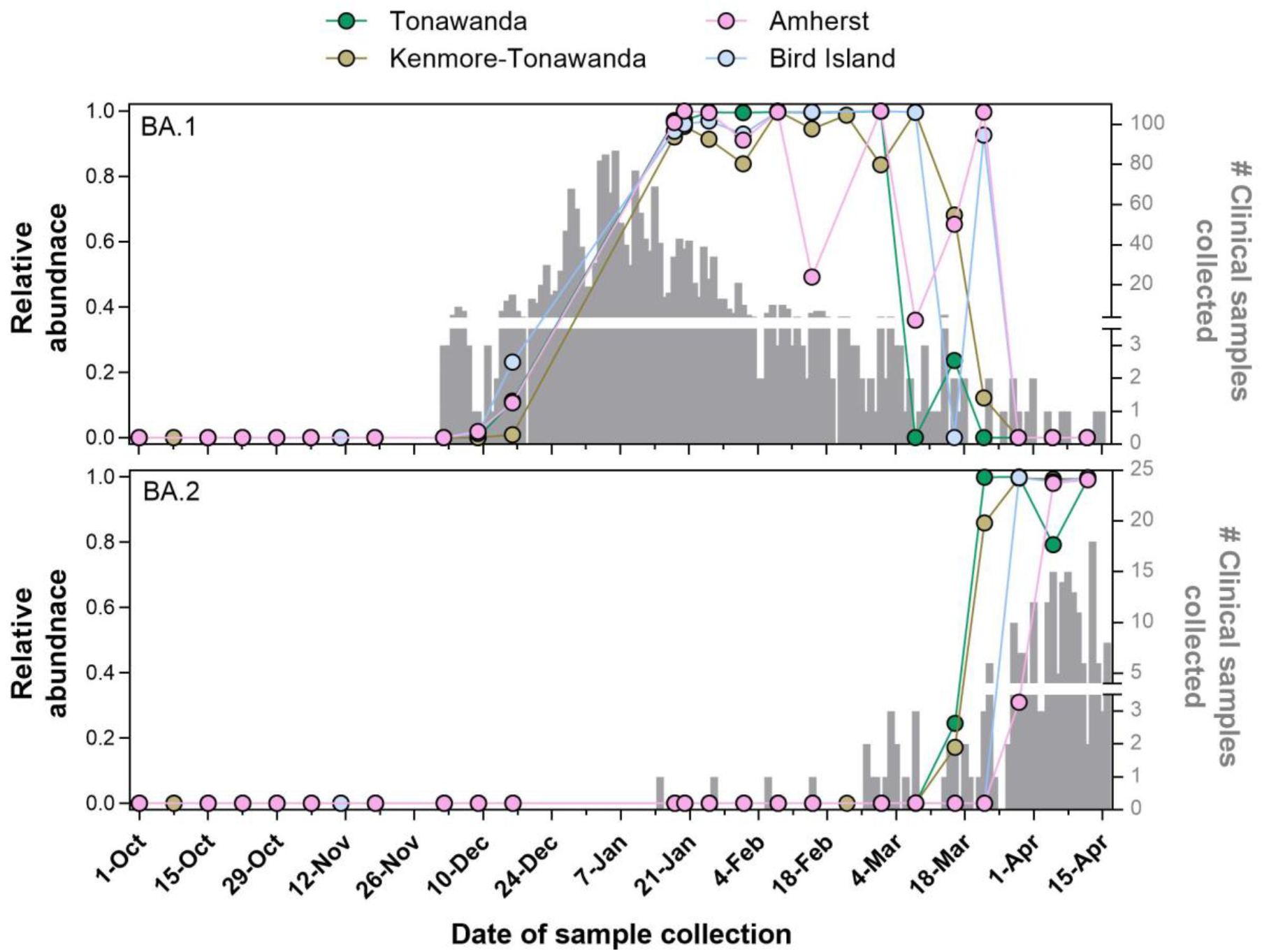
Detection of Omicron BA.1 and BA.2 in wastewater and clinical samples. The relative abundance of Omicron was predicted by Freyja pipeline with the wastewater sequencing data filtered at 50× depth. The relative abundance of BA.1 (top) is the sum of relative abundances of B.1.1.529, BA.1, BA.1.1, and other BA.1.*. The relative abundance of BA.2 (bottom) comprises BA.2.12, BA.2.12.1, and other BA.2.*. The number of clinical samples collected for sequencing was counted on every collection date.

We found that RT-qPCR assays were more sensitive than wastewater WGS for detecting Omicron BA.1-specific mutations, but wastewater WGS was more sensitive for Omicron BA.2-specific mutations (**Table S4**). However, RT-qPCR assays are less sensitive than digital PCR assays,^42^ which were reported to detect variant-specific mutations earlier than genetic sequencing.^9^ Interestingly, our RT-qPCR assays detected S:delH69/V70 mutations (for Omicron BA.1) in the Kenmore-Tonawanda wastewater samples collected in October 2021. Although WGS also detected these mutations, other Omicron-specific mutations in the S gene (Q493R and Q498R) were not detected at the time. Wastewater WGS may thus be more precise than PCR assays for early detection of SARS-CoV-2 variants.

## CONCLUSIONS

Our research demonstrates that tangential-flow filtration to concentrate viruses from wastewater samples enables extraction of nucleic acids of high-enough quality for stable performance of SARS-CoV-2 WGS. Complete or near-complete genomes at a depth of 10× were sequenced from 68% (64/94) of the wastewater samples. Moreover, the tangential-flow filtration method improves the WGS success at lower COVID-19 incidence rates. Our results report a 0.9 probability of WGS success when the COVID-19 incidence rate exceeded 33/100 000 persons. Furthermore, WGS of SARS-CoV-2 in wastewater revealed lineages underrepresented or not detected from patient samples. Future studies are needed to advance wastewater WGS data interpretation and streamline the workflow of wastewater WGS to shorten the turnaround time for early detection.

The developed reusable and likely cost-effective tangential-flow filtration method is readily applicable to other sequencing assays that requires high-quality viral nucleic acids from wastewater. Given the flexibility of concentrating large volumes of wastewater, the tangential-flow filtration system can be further optimized for genomic surveillance of low-abundance viruses in wastewater.

## Data Availability

All data produced in the present work are contained in the manuscript. Raw sequencing data are available in NCBI Sequence Read Archive (SRA) under the BioProject ID: PRJNA877272. Codes for analyzing SARS-CoV-2 lineage in wastewater are available at https://github.com/UBGBC/fastq-to-consensus.

## ASSOCIATED CONTENT

### Supporting information

The supporting information is available free of charge at http://pubs.acs.org.

Inhibition test; RT-qPCR assay details and Limit of detection (LOD) and limit of quantification (LOQ) determination; Details of four sewersheds in this study; Table S2. Details of RT-qPCR assay for quantifying SARS-CoV-2 N gene and S gene mutations; Check list of RT-qPCR experiments according to the MIQE (<u>m</u>inimum information for publication of <u>q</u>uantitative real-time PCR <u>e</u>xperiments) guidelines; Summary of wastewater samples sequenced in this study; Geographic locations of four sewersheds in Erie County, New York; Schematic of tangential-flow filtration system; SARS-CoV-2 N gene levels in wastewater and blank test samples; Determination of limit of detection (LOD; left) and limit of quantification (LOQ; right) of SARS-CoV-2 N gene by RT-qPCR; RT-qPCR inhibition test of SARS-CoV-2 N gene in the wastewater nucleic acid extracts; Comparison of SARS-CoV-2 genome coverage from wastewater with different workflows; Heat map of read depths of SARS-CoV-2 whole-genome sequencing from wastewater samples; Comparison of relative abundances of SARS-CoV-2 lineages estimated by the sequencing data at depths of 10× and 50×; Pair-wise pearson correlation of B.1.2 lineage mutaton spectrum against all other lineage mutation spectra.

## ACKNOWLEDGEMENTS

This work was funded by Erie County Department of Health (ECDOH) award 91287 to Y.Y. and I.M.B. and award 91279 to J.A.S.. We thank undergraduate students Vicky Huang and Danya Hanin for processing the wastewater samples in the lab. We also thank Joseph L. Fiegl for coordinating the wastewater sample collection. The GISAID data used in the manuscript (Identifier: EPI_SET_220906wr) is composed of 4952 individual genome sequences published in GISAID’s EpiCoV database. To view the contributors of each individual sequence with details, visit doi.org/10.55876/gis8.220906wr. Most of the samples (4653/4952) were sequenced at the University at Buffalo. We are grateful for the sequencing work of scientists and healthcare professionals who generated the remaining sequences from the region and made them publicly accessible through GISAID.

## Notes

### Competing Interest Statement

The authors have declared no competing interest.

### Summary of Updates

Authorship is updated.

## REFERENCES

(1) Walensky, R. P.; Walke, H. T.; Fauci, A. S. SARS-CoV-2 variants of concern in the United States-challenges and opportunities. JAMA 2021, 325 (11), 1037–1038. DOI: 10.1001/jama.2021.2294.

(2) Oude Munnink, B. B.; Nieuwenhuijse, D. F.; Stein, M.; O’Toole, A.; Haverkate, M.; Mollers, M.; Kamga, S. K.; Schapendonk, C.; Pronk, M.; Lexmond, P.; et al. Rapid SARS-CoV-2 whole-genome sequencing and analysis for informed public health decision-making in the Netherlands. Nat Med 2020, 26 (9), 1405–1410. DOI: 10.1038/s41591-020-0997-y.

(3) Crits-Christoph, A.; Kantor, R. S.; Olm, M. R.; Whitney, O. N.; Al-Shayeb, B.; Lou, Y. C.; Flamholz, A.; Kennedy, L. C.; Greenwald, H.; Hinkle, A.; et al. Genome Sequencing of Sewage Detects Regionally Prevalent SARS-CoV-2 Variants. mBio 2021, 12 (1), e02703–e02720. DOI: 10.1128/mBio.02703-20.

(4) Nemudryi, A.; Nemudraia, A.; Wiegand, T.; Surya, K.; Buyukyoruk, M.; Cicha, C.; Vanderwood, K. K.; Wilkinson, R.; Wiedenheft, B. Temporal Detection and Phylogenetic Assessment of SARS-CoV-2 in Municipal Wastewater. Cell Rep Med 2020, 1 (6), 100098. DOI: 10.1016/j.xcrm.2020.100098.

(5) Karthikeyan, S.; Levy, J. I.; De Hoff, P.; Humphrey, G.; Birmingham, A.; Jepsen, K.; Farmer, S.; Tubb, H. M.; Valles, T.; Tribelhorn, C. E.; et al. Wastewater sequencing reveals early cryptic SARS-CoV-2 variant transmission. Nature 2022, 609, 101–108. DOI: 10.1038/s41586-022-05049-6.

(6) Smyth, D. S.; Trujillo, M.; Gregory, D. A.; Cheung, K.; Gao, A.; Graham, M.; Guan, Y.; Guldenpfennig, C.; Hoxie, I.; Kannoly, S.; et al. Tracking cryptic SARS-CoV-2 lineages detected in NYC wastewater. Nature Commun 2022, 13 (1), 1–9. DOI: 10.1038/s41467-022-28246-3.

(7) Sims, D.; Sudbery, I.; Ilott, N. E.; Heger, A.; Ponting, C. P. Sequencing depth and coverage: key considerations in genomic analyses. Nat Rev Genet 2014, 15 (2), 121–132. DOI: 10.1038/nrg3642.

(8) Valieris, R.; Drummond, R. D.; Defelicibus, A.; Dias-Neto, E.; Rosales, R. A.; Tojal da Silva, I. A mixture model for determining SARS-Cov-2 variant composition in pooled samples. Bioinformatics 2022, 38 (7), 1809–1815. DOI: 10.1093/bioinformatics/btac047.

(9) Lou, E. G.; Sapoval, N.; McCall, C.; Bauhs, L.; Carlson-Stadler, R.; Kalvapalle, P.; Lai, Y.; Palmer, K.; Penn, R.; Rich, W.; et al. Direct comparison of RT-ddPCR and targeted amplicon sequencing for SARS-CoV-2 mutation monitoring in wastewater. Sci Total Environ 2022, 155059. DOI: 10.1016/j.scitotenv.2022.155059.

(10) Graham, K. E.; Loeb, S. K.; Wolfe, M. K.; Catoe, D.; Sinnott-Armstrong, N.; Kim, S.; Yamahara, K. M.; Sassoubre, L. M.; Grijalva, L. M. M.; Roldan-Hernandez, L.; et al. SARS-CoV-2 RNA in wastewater settled solids is associated with COVID-19 cases in a large urban sewershed. Environ Sci Technol 2021, 55 (1), 488–498. DOI: 10.1021/acs.est.0c06191.

(11) LaTurner, Z. W.; Zong, D. M.; Kalvapalle, P.; Gamas, K. R.; Terwilliger, A.; Crosby, T.; Ali, P.; Avadhanula, V.; Santos, H. H.; Weesner, K.; et al. Evaluating recovery, cost, and throughput of different concentration methods for SARS-CoV-2 wastewater-based epidemiology. Water Res 2021, 197, 117043. DOI: 10.1016/j.watres.2021.117043.

(12) Zheng, X.; Deng, Y.; Xu, X.; Li, S.; Zhang, Y.; Ding, J.; On, H. Y.; Lai, J. C. C.; In Yau, C.; Chin, A. W. H.; et al. Comparison of virus concentration methods and RNA extraction methods for SARS-CoV-2 wastewater surveillance. Sci Total Environ 2022, 824, 153687. DOI: 10.1016/j.scitotenv.2022.153687.

(13) Peccia, J.; Zulli, A.; Brackney, D. E.; Grubaugh, N. D.; Kaplan, E. H.; Casanovas-Massana, A.; Ko, A. I.; Malik, A. A.; Wang, D.; Wang, M.; et al. Measurement of SARS-CoV-2 RNA in wastewater tracks community infection dynamics. Nat Biotechnol 2020, 38 (10), 1164–1167. DOI: 10.1038/s41587-020-0684-z.

(14) Perez-Cataluna, A.; Chiner-Oms, A.; Cuevas-Ferrando, E.; Diaz-Reolid, A.; Falco, I.; Randazzo, W.; Giron-Guzman, I.; Allende, A.; Bracho, M. A.; Comas, I.; et al. Spatial and temporal distribution of SARS-CoV-2 diversity circulating in wastewater. Water Res 2022, 211, 118007. DOI: 10.1016/j.watres.2021.118007.

(15) Fontenele, R. S.; Kraberger, S.; Hadfield, J.; Driver, E. M.; Bowes, D.; Holland, L. A.; Faleye, T. O. C.; Adhikari, S.; Kumar, R.; Inchausti, R.; et al. High-throughput sequencing of SARS-CoV-2 in wastewater provides insights into circulating variants. Water Res 2021, 205, 117710. DOI: 10.1016/j.watres.2021.117710.

(16) Bar-Or, I.; Weil, M.; Indenbaum, V.; Bucris, E.; Bar-Ilan, D.; Elul, M.; Levi, N.; Aguvaev, I.; Cohen, Z.; Shirazi, R.; et al. Detection of SARS-CoV-2 variants by genomic analysis of wastewater samples in Israel. Sci Total Environ 2021, 789, 148002. DOI: 10.1016/j.scitotenv.2021.148002.

(17) Izquierdo-Lara, R.; Elsinga, G.; Heijnen, L.; Munnink, B. B. O.; Schapendonk, C. M. E.; Nieuwenhuijse, D.; Kon, M.; Lu, L.; Aarestrup, F. M.; Lycett, S.; et al. Monitoring SARS-CoV-2 circulation and diversity through community wastewater sequencing, the Netherlands and Belgium. Emerg Infect Dis 2021, 27 (5), 1405–1415. DOI: 10.3201/eid2705.204410.

(18) Lu, X.; Wang, L.; Sakthivel, S. K.; Whitaker, B.; Murray, J.; Kamili, S.; Lynch, B.; Malapati, L.; Burke, S. A.; Harcourt, J.; et al. US CDC real-time reverse transcription PCR panel for detection of Severe Acute Respiratory Syndrome Coronavirus 2. Emerg Infect Dis 2020, 26 (8), 1654. DOI: 10.3201/eid2608.201246.

(19) Lee, W. L.; Gu, X.; Armas, F.; Wu, F.; Chandra, F.; Chen, H.; Xiao, A.; Leifels, M.; Chua, F. J. D.; Kwok, G. W. C.; et al. Quantitative detection of SARS-CoV-2 Omicron BA.1 and BA.2 variants in wastewater through allele-specific RT-qPCR. medRxiv 2022. DOI: 10.1101/2021.12.21.21268077.

(20) Lee, W. L.; Imakaev, M.; Armas, F.; McElroy, K. A.; Gu, X. Q.; Duvallet, C.; Chandra, F.; Chen, H. J.; Leifels, M.; Mendola, S.; et al. Quantitative SARS-CoV-2 Alpha variant B.1.1.7 tracking in wastewater by allele-specific RT-qPCR. Environ Sci Technol Lett 2021, 8 (8), 675–682. DOI: 10.1021/acs.estlett.1c00375.

(21) He, H.; Zhou, P.; Shimabuku, K. K.; Fang, X.; Li, S.; Lee, Y.; Dodd, M. C. Degradation and deactivation of bacterial antibiotic resistance genes during exposure to free chlorine, monochloramine, chlorine dioxide, ozone, ultraviolet light, and hydroxyl radical. Environ Sci Technol 2019, 53 (4), 2013–2026. DOI: 10.1021/acs.est.8b04393.

(22) Bustin, S. A.; Benes, V.; Garson, J. A.; Hellemans, J.; Huggett, J.; Kubista, M.; Mueller, R.; Nolan, T.; Pfaffl, M. W.; Shipley, G. L.; et al. The MIQE guidelines: minimum information for publication of quantitative real-time PCR experiments. Clin Chem 2009, 55 (4), 611–622. DOI: 10.1373/clinchem.2008.112797.

(23) nCoV-2019 sequencing protocol V.1. 2020. https://www.protocols.io/view/ncov-2019-sequencing-protocol-bp2l6n26rgqe/v1?version_warning=no (accessed 2020-01-22).

(24) Wingett, S. W.; Andrews, S. FastQ Screen: A tool for multi-genome mapping and quality control. F1000Research 2018, 7, 1338. DOI: 10.12688/f1000research.15931.2.

(25) Li, H.; Durbin, R. Fast and accurate short read alignment with Burrows-Wheeler transform. Bioinformatics 2009, 25 (14), 1754–1760. DOI: 10.1093/bioinformatics/btp324.

(26) Danecek, P.; Bonfield, J. K.; Liddle, J.; Marshall, J.; Ohan, V.; Pollard, M. O.; Whitwham, A.; Keane, T.; McCarthy, S. A.; Davies, R. M.; et al. Twelve years of SAMtools and BCFtools. Gigascience 2021, 10 (2). DOI: 10.1093/gigascience/giab008 From NLM Medline.

(27) Erie County SARS-CoV-2 wastewater monitoring dashboard. https://erieny.maps.arcgis.com/apps/dashboards/a95853269eec489ea59e5b71571f2e76 (accessed 2022-04-15).

(28) Elbe, S.; Buckland-Merrett, G. Data, disease and diplomacy: GISAID’s innovative contribution to global health. Glob Chall 2017, 1 (1), 33–46. DOI: 10.1002/gch2.1018.

(29) Lamb, N. A., Bard, J.E., Pohlman, A., Boccolucci, A., Yergeau, D.A., Marzullo, B.J., Pope, C., Burstein, G., Tomaszewski, J., Nowak, N.J. and Surtees, J.A. Genomic Surveillance of SARS-CoV-2 in Erie County, New York. medRxiv 2021. DOI: 10.1101/2021.07.01.21259869.

(30) Wickham, H. ggplot2: Elegant Graphics for Data Analysis; Springer-Verlag New York, 2016.

(31) Team, R. C. RStudio: Integrated Development for R. RStudio; PBC, Boston, MA, 2020.

(32) Jahn, K.; Dreifuss, D.; Topolsky, I.; Kull, A.; Ganesanandamoorthy, P.; Fernandez-Cassi, X.; Bänziger, C.; Devaux, A. J.; Stachler, E.; Caduff, L.; et al. Early detection and surveillance of SARS-CoV-2 genomic variants in wastewater using COJAC. Nat Microbiol 2022, 1–10. DOI: 10.1038/s41564-022-01185-x.

(33) Lambisia, A. W.; Mohammed, K. S.; Makori, T. O.; Ndwiga, L.; Mburu, M. W.; Morobe, J. M.; Moraa, E. O.; Musyoki, J.; Murunga, N.; Mwangi, J. N.; et al. Optimization of the SARS-CoV-2 ARTIC Network V4 Primers and Whole Genome Sequencing Protocol. Front Med (Lausanne) 2022, 9, 836728. DOI: 10.3389/fmed.2022.836728.

(34) Clark, C. R.; Hardison, M. T.; Houdeshell, H. N.; Vest, A. C.; Whitlock, D. A.; Skola, D. D.; Koble, J. S.; Oberholzer, M.; Schroth, G. P. Evaluation of an optimized protocol and Illumina ARTIC V4 primer pool for sequencing of SARS-CoV-2 using COVIDSeq™ and DRAGEN™ COVID Lineage App workflow. bioRxiv 2022. DOI: 10.1101/2022.01.07.475443.

(35) Mantilla-Calderon, D.; Huang, K. Y.; Li, A. J.; Chibwe, K.; Yu, X. Q.; Ye, Y.; Liu, L.; Ling, F. Q. Emerging investigator series: meta-analyses on SARS-CoV-2 viral RNA levels in wastewater and their correlations to epidemiological indicators. Environ Sci: Wat Res Technol 2022, 8 (7), 1391–1407. DOI: 10.1039/d2ew00084a.

(36) Itokawa, K.; Sekizuka, T.; Hashino, M.; Tanaka, R.; Kuroda, M. Disentangling primer interactions improves SARS-CoV-2 genome sequencing by multiplex tiling PCR. PLoS One 2020, 15 (9), e0239403. DOI: 10.1371/journal.pone.0239403.

(37) Agrawal, S.; Orschler, L.; Lackner, S. Metatranscriptomic analysis reveals SARS-CoV-2 mutations in wastewater of the Frankfurt metropolitan area in southern Germany. Microbiol Resour Announc 2021, 10 (15), e00280–00221. DOI: 10.1128/MRA.00280-21.

(38) Grant, S. B.; Rekhi, N. V.; Pise, N. R.; Reeves, R. L.; Matsumoto, M.; Wistrom, A.; Moussa, L.; Bay, S.; Kayhanian, M. A review of the contaminants and toxicity associated with particles in stormwater runoff. Terminology 2003, 2 (2), 1–173.

(39) Schrader, C.; Schielke, A.; Ellerbroek, L.; Johne, R. PCR inhibitors - occurrence, properties and removal. J Appl Microbiol 2012, 113 (5), 1014–1026. DOI: 10.1111/j.1365-2672.2012.05384.x.

(40) Tiwari, A.; Lipponen, A.; Hokajarvi, A. M.; Luomala, O.; Sarekoski, A.; Rytkonen, A.; Osterlund, P.; Al-Hello, H.; Juutinen, A.; Miettinen, I. T.; et al. Detection and quantification of SARS-CoV-2 RNA in wastewater influent in relation to reported COVID-19 incidence in Finland. Water Res 2022, 215, 118220. DOI: 10.1016/j.watres.2022.118220.

(41) Larsen, D. A.; Wigginton, K. R. Tracking COVID-19 with wastewater. Nat Biotechnol 2020, 38 (10), 1151–1153. DOI: 10.1038/s41587-020-0690-1.

(42) Ahmed, W., Smith, W.J., Metcalfe, S., Jackson, G., Choi, P.M., Morrison, M., Field, D., Gyawali, P., Bivins, A., Bibby, K. and Simpson, S.L. Comparison of RT-qPCR and RT-dPCR platforms for the trace detection of SARS-CoV-2 RNA in wastewater. ACS ES&T Water 2022. DOI: 10.1021/acsestwater.1c00387.

