## Supporting information for "Improved Robustness of SARS-CoV-2 Whole-Genome Sequencing from Wastewater with a Nonselective Virus Concentration Method"

---

**Contents**

Text S1: Inhibition test S3

Text S2: RT-qPCR assay details and Limit of detection (LOD) and limit of quantification (LOQ) determination S3

Table S1. Details of four sewersheds in this study S6

Table S2. Details of RT-qPCR assay for quantifying SARS-CoV-2 N gene and S gene mutations S7

Table S3. Check list of RT-qPCR experiments according to the MIQE (minimum information for publication of quantitative real-time PCR experiments) guidelines S8

Table S4. Summary of wastewater samples sequenced in this study S9

Figure S1. Geographic locations of four sewersheds in Erie County, New York S11

Figure S2. Schematic of tangential-flow filtration system S12

Figure S3. SARS-CoV-2 N gene levels in wastewater and blank test samples S13

Figure S4. Determination of limit of detection (LOD; left) and limit of quantification (LOQ; right) of SARS-CoV-2 N gene by RT-qPCR S14

Figure S5. RT-qPCR inhibition test of SARS-CoV-2 N gene in the wastewater nucleic acid extracts S15

Figure S6. Comparison of SARS-CoV-2 genome coverage from wastewater with different workflows S16

Figure S7. Heat map of read depths of SARS-CoV-2 whole-genome sequencing from wastewater samples S17

Figure S8. Comparison of relative abundances of SARS-CoV-2 lineages estimated by the sequencing data at depths of 10× and 50× S18

Figure S9. Pair-wise pearson correlation of B.1.2 lineage mutaton spectrum against all other lineage mutation spectra S19

**Text S1: Inhibition test**

We noticed there was some inhibition of RT-qPCR using the wastewater nucleic acid extracts. We tested whether sample dilution would mitigate inhibition and determined the dilution factor needed to remove the inhibitory effects. The SARS-CoV-2 positive control (2019-nCoV research use only kit; IDT) was prepared to a final concentration of $10^3$ gene copies (gc)/μL in nuclease-free water or in wastewater nucleic acid extracts, which had been diluted 5-fold, 10-fold, and 20-fold in nuclease-free water. The $10^3$ gc/μL concentration was ~100× higher than the background SARS-CoV-2 gene concentration in the tested nucleic acid extracts. The SARS-CoV-2 N gene was then measured by RT-qPCR according to the CDC N2 assay.[1] The RT-qPCR was performed with three replicates of the SARS-CoV-2 gene in nuclease-free water and with seven replicates of the SARS-CoV-2 gene prepared in seven different nucleic acid extracts from different wastewater samples. The results suggest that dilution can remove inhibition. The 5-fold dilution decreased inhibition to various levels in different samples (**Figure S5**). The 10-fold and 20-fold dilution removed inhibition across different samples (**Figure S5**). We therefore used the 10-fold dilution to quantify the SARS-CoV-2 N gene, but we used the 5-fold or 10-fold dilution to determine the presence or absence of specific genetic mutations in the SARS-CoV-2 S gene.

**Text S2: RT-qPCR assay details and Limit of detection (LOD) and limit of quantification (LOQ) determination**

*RT-qPCR assays for the SARS-CoV-2 N gene*

The 10-μL RT-qPCR reaction mixtures consisted of 5 μL of 2× iTaq universal probes reaction mix (Bio-Rad), 0.25 μL of 50× iScript reverse transcriptase (Bio-Rad), 0.75 μL of 2019-nCoV_N2 (2019-nCoV research use only kit; IDT), and 4 μL of 10-fold diluted nucleic acid extract. The reactions were heated at 50 °C for 15 min, 95°C for 1 min, and 40 cycles of 95 °C for 10 s and 60 °C for 30 s. The standard curve of SARS-CoV-2 N gene had an average $R^2$ of >0.98 and average efficiency of 1.13 (**Table S2**).

*RT-qPCR assays for genetic mutations in the SARS-CoV-2 S gene*

Each assay was optimized with Twist synthetic RNA controls (TWIST Bioscience) for optimal amplification (**Table S2**). The S gene assays were only used to measure the presence and absence of specific S gene mutations. Specifically, for the Delta assay that targets WT493-498 and the generic Omicron assay that targets Q493R and Q498R, the 20-μL reaction mixtures contained 5 μL of Reliance one-step multiplex RT-qPCR supermix (Bio-Rad), 1.2 μL of forward primers at 10 μM, 1.2 μL of reverse primers at 10 μM, 0.5 μL of probe at 10 μM, 10 μL of 5-fold diluted nucleic acid extract, and 2.1 μL of nuclease-free water. The reactions were conducted under the following thermocycling conditions: 50 °C for 10 min, 95°C for 10 min, and 45 cycles of 95 °C for 10 s and 60 °C for 30 s. For the Omicron BA.1 assay that measures delL24/P25/P26 and A27S, the 20-μL reaction mixtures consisted of 5 μL of reliance one-step multiplex RT-qPCR supermix, 1.2 μL of forward primers at 10 μM, 1.2 μL of reverse primers at 10 μM, 0.4 μL of probe at 10 μM, 10 μL of 10-fold diluted nucleic acid extract, and 2.2 μL of nuclease-free water. The following thermocycling conditions were used: 50 °C for 10 min, 95°C for 10 min, and 45 cycles of 95 °C for 10 s and 60 °C for 30 s. For the Omicron BA.2 assay that targets delH69/V70, the 20-μL reaction mixtures contained 5 μL of Reliance one-step multiplex

RT-qPCR supermix, 1.2 µL of forward primers at 10 µM, 1.2 µL of reverse primers at 10 µM, 0.5 µL of probe at 10 µM, 10 µL of 10-fold diluted nucleic acid extract, and 2.1 µL of nuclease-free water. The reactions were conducted under the following thermocycling conditions: 50 °C for 10 min, 95°C for 10 min, and 45 cycles of 95 °C for 10 s and 55 °C for 30 s.

*LOD and LOQ determination for the SARS-CoV-2 N gene*

The limit of detection (LOD) and limit of quantification (LOQ) of the SARS-CoV-2 N gene with the CDC N2 RT-qPCR assay were determined according to a modified procedure described by He et al.[2] Specifically, different concentrations of SARS-CoV-2 positive control (2019-nCoV research use only kit; IDT) were prepared as $10^4$, $10^3$, $10^2$, $10^1$, 5, 2, 1, and 0.5 gc/µL in nuclease-free water. We performed four PCR replicates of the $10^4$, $10^3$, $10^2$, and $10^1$ gc/µL standards and eight PCR replicates of the 5, 2, 1, and 0.5 gc/µL standards. The resulting detection rates of each standard based on their $C_T$ values (<40) were fitted with a nonlinear regression model. The LOD was determined as the lowest concentration at which the detection rate was >95%. The LOQ was determined as the lowest concentration at which the coefficient of variation (CV) was <40%. The CV of each standard was calculated according to the following equation:

$$CV = 100\% \sqrt{(1+E)^{(SDct)^2 \ln(1+E)} - 1}.$$

Here, SDct is the standard deviation of $C_T$ values measured for each standard concentration; E is the amplification efficiency determined using the slope of standard curve within the linear range, $E = 10^{1/slope}$.

As a result, the LOD was determined to be 1.8 gc/µL, and the LOQ was determined to be 5 gc/µL in the nucleic acid extracts (**Figure S4**).

**Table S1.** Details of four sewersheds in this study

| **Wastewater treatment plant** | **Sewershed** | **Estimated population** | **Permit flow rate (MGD*)** | **Composite sampling type** | **Sewer type** |
|---|---|---|---|---|---|
| Tonawanda wastewater treatment plant | Tonawanda | 14,873 | 2.5-5 | Flow weighted | Separate /Combined |
| | Kenmore-Tonawanda | 70,470 | 25 | Flow weighted | Separate |
| Amherst wastewater treatment plant | Amherst | 140,324 | 48 | Time weighted | Separate |
| Bird Island wastewater treatment plant | Bird Island | 437,357 | 180 | Time weighted | Combined |

*MGD = millions of gallons per day

**Table S2.** Details of RT-qPCR assay for quantifying SARS-CoV-2 N gene and S gene mutations

| Assay name | Target gene region or mutations | Forward Primer (5' to 3', F)<br>Reverse Primer (5' to 3', R)<br>Probe (5' to 3', P) | Size of amplicon (bp) | Source of control/standard | Calibration curve slop (intercept) | Efficiency (%) | $R^2$ | Ref. |
|---|---|---|---|---|---|---|---|---|
| CDC-N2 | N gene | TTA CAA ACA TTG GCC GCA AA (F)<br>GCG CGA CAT TCC GAA GAA (R)<br>6-FAM-ACA ATT TGC /ZEN/ CCC CAG CGC TTC AG-IABkFQ (P) | 67 | 2019-nCoV research use only kit (IDT) | -3.057 (+36.69) | 113 | 0.984 | [1] |
| Delta | S: Q483, Q498 | CTT TCC TTT ACA ATC ATA TGG TTT CCA (F)<br>AGT TGC TGG TGC ATG TAG AA (R)<br>6-FAM-ACC CAC TWA /ZEN/ TGG TGT TGG TYA CCA-IABkFQ (P) | 103 | Twist synthetic RNA control 23 (B.1.617.2) (TWIST Bioscience) | | | | [3] |
| Omicron | S: Q483R, Q498R | CTT TCC TTT ACG ATC ATA TAG TTT CCG (F)<br>AGT TGC TGG TGC ATG TAG AA (R)<br>6-FAM-ACC CAC TWA /ZEN/ TGG TGT TGG TYA CCA-IABkFQ (P) | 103 | Twist synthetic RNA control 48 (B.1.1.529/BA.1) (TWIST Bioscience) | | | | [3] |
| Omicron BA.1 | S: delH69/V70 | ATG TTA CTT GGT TCC ATG CTA TCT C (F)<br>AAA TGG TAG GAC AGG GTT ATC AA (R)<br>6-FAM-TCT CTG GGA /ZEN/ CCA ATG GTA CTA AGA GGT-IABkFQ (P) | 77 | Twist synthetic RNA control 48 (B.1.1.529/BA.1) (TWIST Bioscience) | | | | [3, 4] |
| Omicron BA.2 | S: delL24, delP25, delP26, A27S | TGT TAA TCT TAT AAC CAG AAC TCA ATC ATA (F)<br>AGA ACA AGT CCT GAG TTG AAT GTA (R)<br>6-FAM-TCA CAC GTG /ZEN/ GTG TTT ATT ACC CTG ACA-IABkFQ (P) | 113 | Twist synthetic RNA control 50 (B.1.1.529/BA.2) (TWIST Bioscience) | | | | [3] |

**Table S3.** Check list of RT-qPCR experiments according to the MIQE (minimum information for publication of quantitative real-time PCR experiments) guidelines.[5]

**Table S4.** Summary of wastewater samples sequenced in this study

| | | | | | | | Sequencing, | | | | RT-qPCR, Ct value (**) | | | |
|---|---|---|---|---|---|---|---|---|---|---|---|---|---|---|
| SeqID | seqName | Collection date | Location | COVID-19 incidence rate (per 100,000) | RT-qPCR, Ct (SARS-CoV-2 N) | Genome coverage at 10x depth | WT 493-498 (Delta) | S:Q493R, S:Q498R (Omicron) | S: delH69, S: delV70 (Omicron BA.1) | S:delL24, S:delP25, S:delP26, S:A27S (Omicron BA.2) | WT 493-498 (Delta) | S:Q493R, S:Q498R (Omicron) | S: delH69, S: delV70 (Omicron BA.1) | S:delL24, S:delP25, S:delP26, S:A27S (Omicron BA.2) |
| UF_10_01 | hCov-19/USA/NY-UF_10_01/2021 | 10/1/2021 | Amherst | 18.80 | 33.7 | 97.8 | + | - | - | - | | | | |
| UF_10_12 | hCov-19/USA/NY-UF_10_12/2021 | 10/15/2021 | Amherst | 17.30 | 31.4 | 81.8 | + | - | - | - | | | | |
| UF_10_19 | hCov-19/USA/NY-UF_10_19/2021 | 10/22/2021 | Amherst | 22.00 | 32.2 | 97.4 | + | - | - | - | | | | |
| UF_10_25 | hCov-19/USA/NY-UF_10_25/2021 | 10/29/2021 | Amherst | 27.20 | NaN * | 91.3 | + | - | - | - | | | | |
| UF_11_01 | hCov-19/USA/NY-UF_11_01/2021 | 11/5/2021 | Amherst | 33.00 | 32.3 | 83.9 | + | - | - | - | 34.7 | NaN | | |
| UF_11_10 | hCov-19/USA/NY-UF_11_10/2021 | 11/18/2021 | Amherst | 52.00 | 32.9 | 99.8 | + | - | - | - | 35.4 | NaN | | |
| UF_12_01 | hCov-19/USA/NY-UF_12_01/2022 | 12/2/2021 | Amherst | 60.00 | 31.2 | 99.8 | + | - | - | - | 33.2 | 39.3 | 38.4 | |
| UF_12_05 | hCov-19/USA/NY-UF_12_05/2022 | 12/9/2021 | Amherst | 41.50 | 31.8 | 99.8 | + | - | - | - | | | 39.1 | |
| UF_12_09 | hCov-19/USA/NY-UF_12_09/2022 | 12/16/2021 | Amherst | 40.20 | 33.0 | 99.3 | - | + | + | - | 35.2 | 37.2 | 37.4 | |
| UF_01_01 | hCov-19/USA/NY-UF_01_01/2022 | 1/18/2022 | Amherst | 101.60 | 31.7 | 99.4 | - | + | + | - | 39.7 | 33.6 | 34.3 | |
| UF_01_05 | hCov-19/USA/NY-UF_01_05/2022 | 1/20/2022 | Amherst | 99.10 | 37.1 | 25.2 | - | - | - | - | 39.3 | 38.6 | NaN | |
| UF_01_09 | hCov-19/USA/NY-UF_01_09/2022 | 1/25/2022 | Amherst | 67.60 | 33.0 | 99.1 | - | + | + | - | 39.9 | 34.3 | 35.4 | |
| UF_02_01 | hCov-19/USA/NY-UF_02_01/2022 | 2/1/2022 | Amherst | 36.20 | 33.9 | 97.1 | - | + | + | - | | | 36.2 | |
| UF_02_05 | hCov-19/USA/NY-UF_02_05/2022 | 2/8/2022 | Amherst | 20.80 | 34.7 | 97.2 | - | + | + | - | | | 37.3 | |
| UF_02_17 | hCov-19/USA/NY-UF_02_17/2022 | 2/15/2022 | Amherst | 9.30 | 35.8 | 95.1 | + | - | + | - | | | 37.7 | NaN |
| UF_02_21 | hCov-19/USA/NY-UF_02_21/2022 | 3/1/2022 | Amherst | 9.90 | 34.4 | 30.4 | + | - | - | - | | 38.5 | 39.7 | NaN |
| UF_03_08 | hCov-19/USA/NY-UF_03_08/2022 | 3/8/2022 | Amherst | 5.60 | 36.6 | 15.0 | + | - | - | - | | 39.4 | 39.7 | NaN |
| UF_03_02 | hCov-19/USA/NY-UF_03_02/2022 | 3/16/2022 | Amherst | 6.40 | NaN | 30.5 | + | - | - | - | | 39.4 | NaN | NaN |
| UF_03_12 | hCov-19/USA/NY-UF_03_12/2022 | 3/22/2022 | Amherst | 10.03 | 35.1 | 62.7 | - | + | - | - | | 36.5 | 38.5 | NaN |
| UF_03_16 | hCov-19/USA/NY-UF_03_16/2022 | 3/29/2022 | Amherst | 15.68 | 34.1 | 90.6 | - | + | - | + | | 35.0 | 38.9 | NaN |
| UF_04_01 | hCov-19/USA/NY-UF_04_01/2022 | 4/5/2022 | Amherst | 18.80 | 35.5 | 61.3 | + | - | - | + | | | NaN | 39.7 |
| UF_04_05 | hCov-19/USA/NY-UF_04_05/2022 | 4/12/2022 | Amherst | 16.00 | 36.8 | 97.1 | - | + | - | + | | | NaN | 37.8 |
| UF_10_04 | hCov-19/USA/NY-UF_10_04/2021 | 10/1/2021 | Bird Island | 17.40 | 34.1 | 91.0 | + | - | - | - | | | | |
| UF_10_15 | hCov-19/USA/NY-UF_10_15/2021 | 10/15/2021 | Bird Island | 20.50 | 31.7 | 77.8 | + | - | - | - | | | | |
| UF_10_22 | hCov-19/USA/NY-UF_10_22/2021 | 10/22/2021 | Bird Island | 22.00 | 33.0 | 90.8 | + | - | - | - | | | | |
| UF_10_28 | hCov-19/USA/NY-UF_10_28/2021 | 10/29/2021 | Bird Island | 25.20 | 34.9 | 97.1 | + | - | - | - | | | | |
| UF_11_04 | hCov-19/USA/NY-UF_11_04/2021 | 11/5/2021 | Bird Island | 36.00 | 33.3 | 60.6 | + | - | - | - | 35.1 | NaN | | |
| UF_11_07 | hCov-19/USA/NY-UF_11_07/2021 | 11/11/2021 | Bird Island | 42.60 | 32.3 | 99.8 | + | - | - | - | | | | |
| UF_11_13 | hCov-19/USA/NY-UF_11_13/2021 | 11/18/2021 | Bird Island | 50.70 | 38.9 | 99.8 | + | - | - | - | 35.5 | NaN | | |
| UF_12_04 | hCov-19/USA/NY-UF_12_04/2022 | 12/2/2021 | Bird Island | 60.20 | 31.4 | 99.8 | + | - | - | - | 33.7 | 39.8 | NaN | |
| UF_12_08 | hCov-19/USA/NY-UF_12_08/2022 | 12/9/2021 | Bird Island | 47.10 | 31.5 | 99.8 | + | - | - | - | | | 38.7 | |
| UF_01_12 | hCov-19/USA/NY-UF_01_12/2022 | 12/16/2021 | Bird Island | 57.00 | 32.6 | 99.8 | - | + | - | - | 39.8 | 35.6 | | |
| UF_01_04 | hCov-19/USA/NY-UF_01_04/2022 | 1/18/2022 | Bird Island | 97.00 | 33.1 | 98.6 | - | + | + | - | 39.0 | 34.8 | 35.5 | |
| UF_01_08 | hCov-19/USA/NY-UF_01_08/2022 | 1/20/2022 | Bird Island | 87.80 | 31.9 | 99.1 | - | + | + | - | 38.6 | 33.8 | 35.2 | |
| UF_12_12 | hCov-19/USA/NY-UF_12_12/2022 | 1/25/2022 | Bird Island | 44.30 | 32.0 | 98.0 | - | + | + | - | | | 37.2 | |
| UF_02_04 | hCov-19/USA/NY-UF_02_04/2022 | 2/1/2022 | Bird Island | 30.60 | 33.9 | 97.5 | - | + | + | - | 39.1 | 35.0 | 35.4 | |
| UF_02_08 | hCov-19/USA/NY-UF_02_08/2022 | 2/8/2022 | Bird Island | 19.60 | 35.2 | 97.1 | - | + | + | - | | 36.2 | 37.5 | |
| UF_02_20 | hCov-19/USA/NY-UF_02_20/2022 | 2/15/2022 | Bird Island | 6.80 | 34.1 | 57.0 | + | - | + | - | | | NaN | |
| UF_02_24 | hCov-19/USA/NY-UF_02_24/2022 | 3/1/2022 | Bird Island | 6.50 | 34.0 | 64.2 | - | + | + | - | | 37.8 | 39.1 | NaN |
| UF_03_11 | hCov-19/USA/NY-UF_03_11/2022 | 3/8/2022 | Bird Island | 5.30 | 35.8 | 60.0 | - | + | + | - | | 38.2 | NaN | NaN |
| UF_03_04 | hCov-19/USA/NY-UF_03_04/2022 | 3/16/2022 | Bird Island | 6.20 | NaN | 53.7 | - | + | - | + | | 38.1 | 38.1 | NaN |
| UF_03_15 | hCov-19/USA/NY-UF_03_15/2022 | 3/22/2022 | Bird Island | 8.34 | 37.1 | 58.0 | + | - | + | - | | 35.6 | 38.8 | NaN |
| UF_03_19 | hCov-19/USA/NY-UF_03_19/2022 | 3/29/2022 | Bird Island | 11.28 | 35.2 | 92.2 | - | + | - | + | | 36.2 | 39.6 | NaN |
| UF_04_04 | hCov-19/USA/NY-UF_04_04/2022 | 4/5/2022 | Bird Island | 17.40 | 33.0 | 98.2 | - | + | - | + | | | NaN | 38.1 |
| UF_04_08 | hCov-19/USA/NY-UF_04_08/2022 | 4/12/2022 | Bird Island | 16.60 | 36.7 | 76.0 | + | - | - | + | | | NaN | NaN |
| UF_10_03 | hCov-19/USA/NY-UF_10_03/2021 | 10/1/2021 | Kenmore Tonawanda | 24.50 | 32.3 | 99.0 | + | - | + | - | | | 37.6 | |

| | | | | | | | Sequencing, | | | | RT-qPCR, Ct value (**) | | | |
|---|---|---|---|---|---|---|---|---|---|---|---|---|---|---|
| SeqID | seqName | Collection date | Location | COVID-19 incidence rate (per 100,000) | RT-qPCR, Ct (SARS-CoV-2 N) | Genome coverage at 10x depth | WT 493-498 (Delta) | S:Q493R, S:Q498R (Omicron) | S: delH69, S: delV70 (Omicron BA.1) | S:delL24, S:delP25, S:delP26, S:A27S (Omicron BA.2) | WT 493-498 (Delta) | S:Q493R, S:Q498R (Omicron) | S: delH69, S: delV70 (Omicron BA.1) | S:delL24, S:delP25, S:delP26, S:A27S (Omicron BA.2) |
| UF_10_01 | hCov-19/USA/NY-UF_10_01/2021 | 10/1/2021 | Amherst | 18.80 | 33.7 | 97.8 | + | - | - | - | | | | |
| UF_10_12 | hCov-19/USA/NY-UF_10_12/2021 | 10/15/2021 | Amherst | 17.30 | 31.4 | 81.8 | + | - | - | - | | | | |
| UF_10_19 | hCov-19/USA/NY-UF_10_19/2021 | 10/22/2021 | Amherst | 22.00 | 32.2 | 97.4 | + | - | - | - | | | | |
| UF_10_25 | hCov-19/USA/NY-UF_10_25/2021 | 10/29/2021 | Amherst | 27.20 | NaN * | 91.3 | + | - | - | - | | | | |
| UF_11_01 | hCov-19/USA/NY-UF_11_01/2021 | 11/5/2021 | Amherst | 33.00 | 32.3 | 83.9 | + | - | - | - | 34.7 | NaN | | |
| UF_11_10 | hCov-19/USA/NY-UF_11_10/2021 | 11/18/2021 | Amherst | 52.00 | 32.9 | 99.8 | + | - | - | - | 35.4 | NaN | | |
| UF_12_01 | hCov-19/USA/NY-UF_12_01/2022 | 12/2/2021 | Amherst | 60.00 | 31.2 | 99.8 | + | - | - | - | 33.2 | 39.3 | 38.4 | |
| UF_12_05 | hCov-19/USA/NY-UF_12_05/2022 | 12/9/2021 | Amherst | 41.50 | 31.8 | 99.8 | + | - | - | - | | | 39.1 | |
| UF_12_09 | hCov-19/USA/NY-UF_12_09/2022 | 12/16/2021 | Amherst | 40.20 | 33.0 | 99.3 | - | + | + | - | 35.2 | 37.2 | 37.4 | |
| UF_01_01 | hCov-19/USA/NY-UF_01_01/2022 | 1/18/2022 | Amherst | 101.60 | 31.7 | 99.4 | - | + | + | - | 39.7 | 33.6 | 34.3 | |
| UF_01_05 | hCov-19/USA/NY-UF_01_05/2022 | 1/20/2022 | Amherst | 99.10 | 37.1 | 25.2 | - | - | - | - | 39.3 | 38.6 | NaN | |
| UF_01_09 | hCov-19/USA/NY-UF_01_09/2022 | 1/25/2022 | Amherst | 67.60 | 33.0 | 99.1 | - | + | + | - | 39.9 | 34.3 | 35.4 | |
| UF_02_01 | hCov-19/USA/NY-UF_02_01/2022 | 2/1/2022 | Amherst | 36.20 | 33.9 | 97.1 | - | + | + | - | | | 36.2 | |
| UF_02_05 | hCov-19/USA/NY-UF_02_05/2022 | 2/8/2022 | Amherst | 20.80 | 34.7 | 97.2 | - | + | + | - | | | 37.3 | |
| UF_02_17 | hCov-19/USA/NY-UF_02_17/2022 | 2/15/2022 | Amherst | 9.30 | 35.8 | 95.1 | + | - | + | - | | | 37.7 | NaN |
| UF_02_21 | hCov-19/USA/NY-UF_02_21/2022 | 3/1/2022 | Amherst | 9.90 | 34.4 | 30.4 | + | - | - | - | | 38.5 | 39.7 | NaN |
| UF_03_08 | hCov-19/USA/NY-UF_03_08/2022 | 3/8/2022 | Amherst | 5.60 | 36.6 | 15.0 | + | - | - | - | | 39.4 | 39.7 | NaN |
| UF_03_02 | hCov-19/USA/NY-UF_03_02/2022 | 3/16/2022 | Amherst | 6.40 | NaN | 30.5 | + | - | - | - | | 39.4 | NaN | NaN |
| UF_03_12 | hCov-19/USA/NY-UF_03_12/2022 | 3/22/2022 | Amherst | 10.03 | 35.1 | 62.7 | - | + | - | - | | 36.5 | 38.5 | NaN |
| UF_03_16 | hCov-19/USA/NY-UF_03_16/2022 | 3/29/2022 | Amherst | 15.68 | 34.1 | 90.6 | - | + | - | + | | 35.0 | 38.9 | NaN |
| UF_04_01 | hCov-19/USA/NY-UF_04_01/2022 | 4/5/2022 | Amherst | 18.80 | 35.5 | 61.3 | + | - | - | + | | | NaN | 39.7 |
| UF_04_05 | hCov-19/USA/NY-UF_04_05/2022 | 4/12/2022 | Amherst | 16.00 | 36.8 | 97.1 | - | + | - | + | | | NaN | 37.8 |
| UF_10_04 | hCov-19/USA/NY-UF_10_04/2021 | 10/1/2021 | Bird Island | 17.40 | 34.1 | 91.0 | + | - | - | - | | | | |
| UF_10_15 | hCov-19/USA/NY-UF_10_15/2021 | 10/15/2021 | Bird Island | 20.50 | 31.7 | 77.8 | + | - | - | - | | | | |
| UF_10_22 | hCov-19/USA/NY-UF_10_22/2021 | 10/22/2021 | Bird Island | 22.00 | 33.0 | 90.8 | + | - | - | - | | | | |
| UF_10_28 | hCov-19/USA/NY-UF_10_28/2021 | 10/29/2021 | Bird Island | 25.20 | 34.9 | 97.1 | + | - | - | - | | | | |
| UF_11_04 | hCov-19/USA/NY-UF_11_04/2021 | 11/5/2021 | Bird Island | 36.00 | 33.3 | 60.6 | + | - | - | - | 35.1 | NaN | | |
| UF_11_07 | hCov-19/USA/NY-UF_11_07/2021 | 11/11/2021 | Bird Island | 42.60 | 32.3 | 99.8 | + | - | - | - | | | | |
| UF_11_13 | hCov-19/USA/NY-UF_11_13/2021 | 11/18/2021 | Bird Island | 50.70 | 38.9 | 99.8 | + | - | - | - | 35.5 | NaN | | |
| UF_12_04 | hCov-19/USA/NY-UF_12_04/2022 | 12/2/2021 | Bird Island | 60.20 | 31.4 | 99.8 | + | - | - | - | 33.7 | 39.8 | NaN | |
| UF_12_08 | hCov-19/USA/NY-UF_12_08/2022 | 12/9/2021 | Bird Island | 47.10 | 31.5 | 99.8 | + | - | - | - | | | 38.7 | |
| UF_01_12 | hCov-19/USA/NY-UF_01_12/2022 | 12/16/2021 | Bird Island | 57.00 | 32.6 | 99.8 | - | + | - | - | 39.8 | 35.6 | | |
| UF_01_04 | hCov-19/USA/NY-UF_01_04/2022 | 1/18/2022 | Bird Island | 97.00 | 33.1 | 98.6 | - | + | + | - | 39.0 | 34.8 | 35.5 | |
| UF_01_08 | hCov-19/USA/NY-UF_01_08/2022 | 1/20/2022 | Bird Island | 87.80 | 31.9 | 99.1 | - | + | + | - | 38.6 | 33.8 | 35.2 | |
| UF_12_12 | hCov-19/USA/NY-UF_12_12/2022 | 1/25/2022 | Bird Island | 44.30 | 32.0 | 98.0 | - | + | + | - | | | 37.2 | |
| UF_02_04 | hCov-19/USA/NY-UF_02_04/2022 | 2/1/2022 | Bird Island | 30.60 | 33.9 | 97.5 | - | + | + | - | 39.1 | 35.0 | 35.4 | |
| UF_02_08 | hCov-19/USA/NY-UF_02_08/2022 | 2/8/2022 | Bird Island | 19.60 | 35.2 | 97.1 | - | + | + | - | | 36.2 | 37.5 | |
| UF_02_20 | hCov-19/USA/NY-UF_02_20/2022 | 2/15/2022 | Bird Island | 6.80 | 34.1 | 57.0 | + | - | + | - | | | NaN | |
| UF_02_24 | hCov-19/USA/NY-UF_02_24/2022 | 3/1/2022 | Bird Island | 6.50 | 34.0 | 64.2 | - | + | + | - | | 37.8 | 39.1 | NaN |
| UF_03_11 | hCov-19/USA/NY-UF_03_11/2022 | 3/8/2022 | Bird Island | 5.30 | 35.8 | 60.0 | - | + | + | - | | 38.2 | NaN | NaN |
| UF_03_04 | hCov-19/USA/NY-UF_03_04/2022 | 3/16/2022 | Bird Island | 6.20 | NaN | 53.7 | - | + | - | + | | 38.1 | 38.1 | NaN |
| UF_03_15 | hCov-19/USA/NY-UF_03_15/2022 | 3/22/2022 | Bird Island | 8.34 | 37.1 | 58.0 | + | - | + | - | | 35.6 | 38.8 | NaN |
| UF_03_19 | hCov-19/USA/NY-UF_03_19/2022 | 3/29/2022 | Bird Island | 11.28 | 35.2 | 92.2 | - | + | - | + | | 36.2 | 39.6 | NaN |
| UF_04_04 | hCov-19/USA/NY-UF_04_04/2022 | 4/5/2022 | Bird Island | 17.40 | 33.0 | 98.2 | - | + | - | + | | | NaN | 38.1 |
| UF_04_08 | hCov-19/USA/NY-UF_04_08/2022 | 4/12/2022 | Bird Island | 16.60 | 36.7 | 76.0 | + | - | - | + | | | NaN | NaN |
| UF_10_03 | hCov-19/USA/NY-UF_10_03/2021 | 10/1/2021 | Kenmore Tonawanda | 24.50 | 32.3 | 99.0 | + | - | + | - | | | 37.6 | |

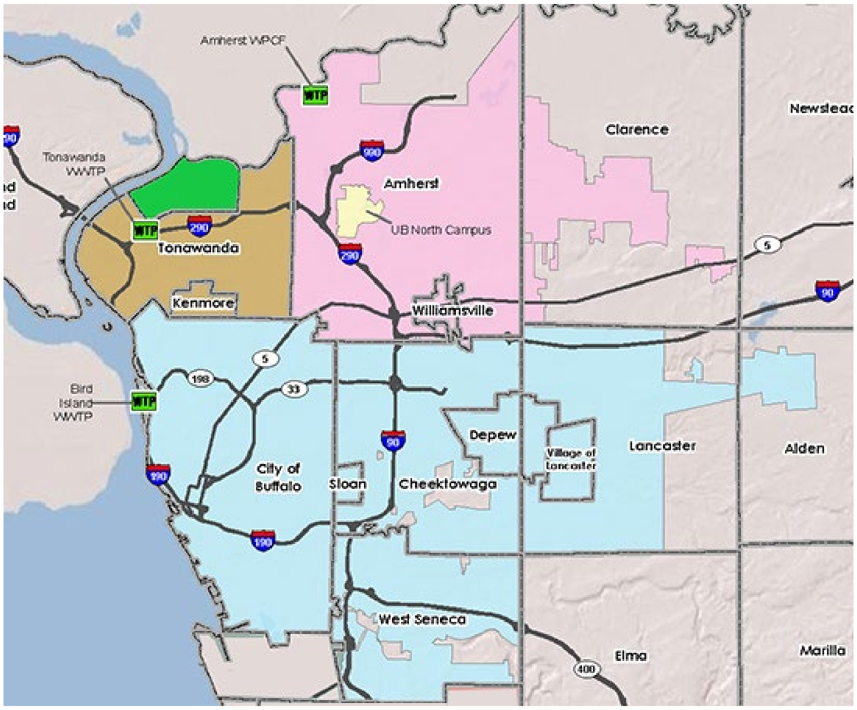


**Figure S1.** Geographic locations of four sewersheds in Erie County, New York: Tonawanda (green), Kenmore-Tonawanda (brown), Amherst (pink), and Bird Island (blue).

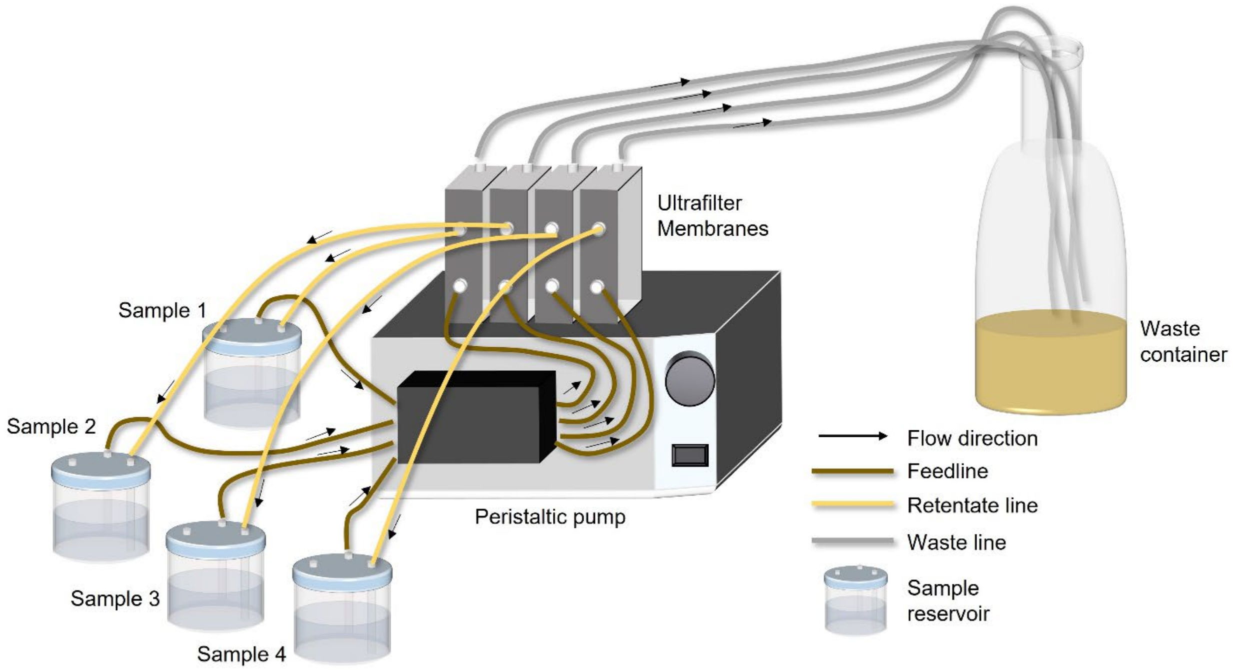


**Figure S2.** Schematic of tangential-flow filtration system. During the operation, 125 mL of solids-removed wastewater samples were loaded from sample reservoirs to 30-kDa ultrafilter membranes at a feedline flow rate of 8.5 mL/min. The membrane filtrate was collected in the waste container.

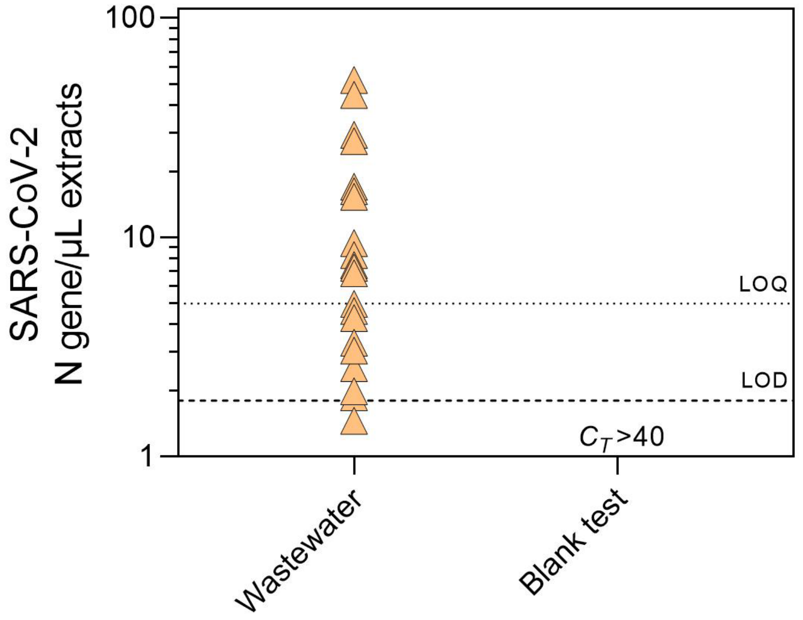


**Figure S3.** SARS-CoV-2 N gene levels in wastewater ($n$ = 24) and blank test samples ($n$ = 24). In the blank test, autoclaved Milli-Q water (125 mL) was concentrated with the same procedures used for wastewater samples after the tangential-flow filtration system was washed with 0.5 M NaOH. The nucleic acids were then extracted from the concentrated Milli-Q water samples, and the concentrations of SARS-CoV-2 N gene were quantified by RT-qPCR according to the CDC N2 assay. No signals were detected in the blank test samples ($C_T$ > 40). The limit of quantification (LOD) and limit of detection (LOD) were determined as described in **Text S2** and **Figure S4**.

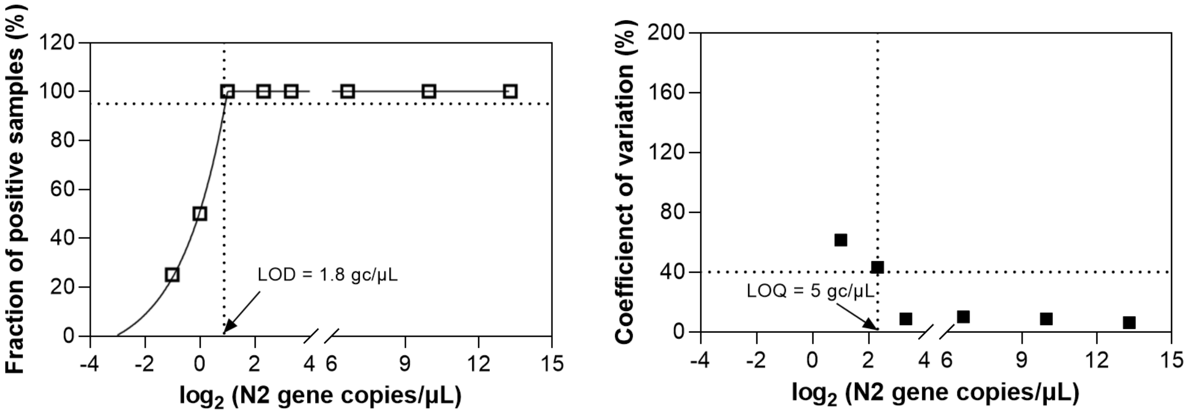


**Figure S4.** Determination of limit of detection (LOD; left) and limit of quantification (LOQ; right) of SARS-CoV-2 N gene by RT-qPCR (CDC N2 assay). The horizontal dashed line in the LOD plot represents 95% positive detection. The horizontal dashed line in the LOQ plot represents 40% of the coefficient of variation.

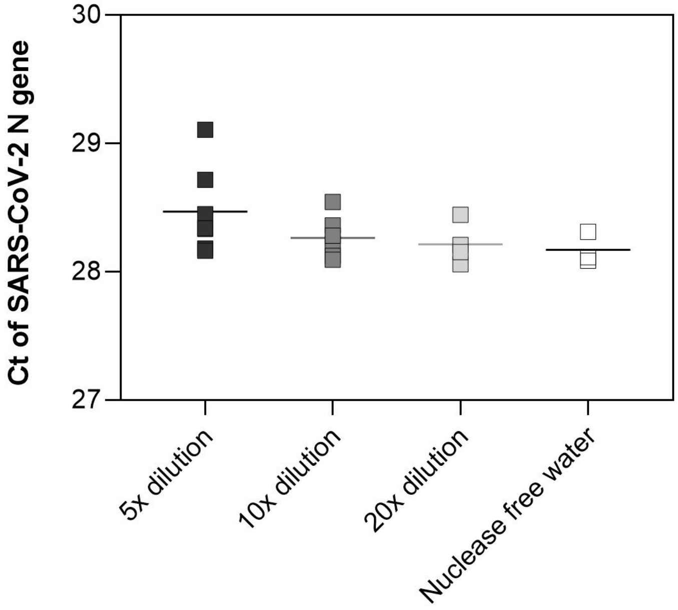


**Figure S5**. RT-qPCR inhibition test of SARS-CoV-2 N gene in the wastewater nucleic acid extracts. SARS-CoV-2 standards ($10^3$ gc/μL) were prepared in wastewater nucleic acid extracts that were 5-fold, 10-fold, and 20-fold diluted (*n* = 7) and in nuclease-free water (*n* = 3). The bars represent mean values.

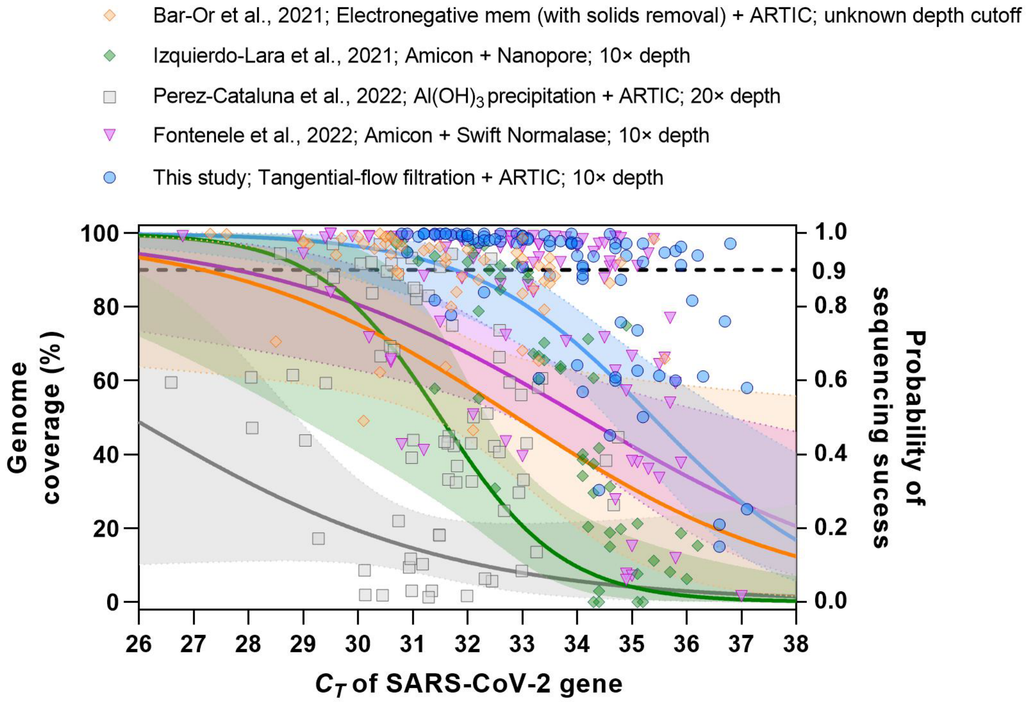


**Figure S6.** Comparison of SARS-CoV-2 genome coverage and probability of sequencing success from wastewater with different workflows.[6-9] The dashed line represents the 0.9 probability of sequencing success. The workflows were selected for comparison according to the following criteria: (i) used a nonselective method to concentrate viruses from wastewater; (ii) processed >50 wastewater samples (excluded a proof-of-concept study); (iii) reported the $C_T$ values of SARS-CoV-2 gene and the corresponding genome coverage in the main text or supporting information.

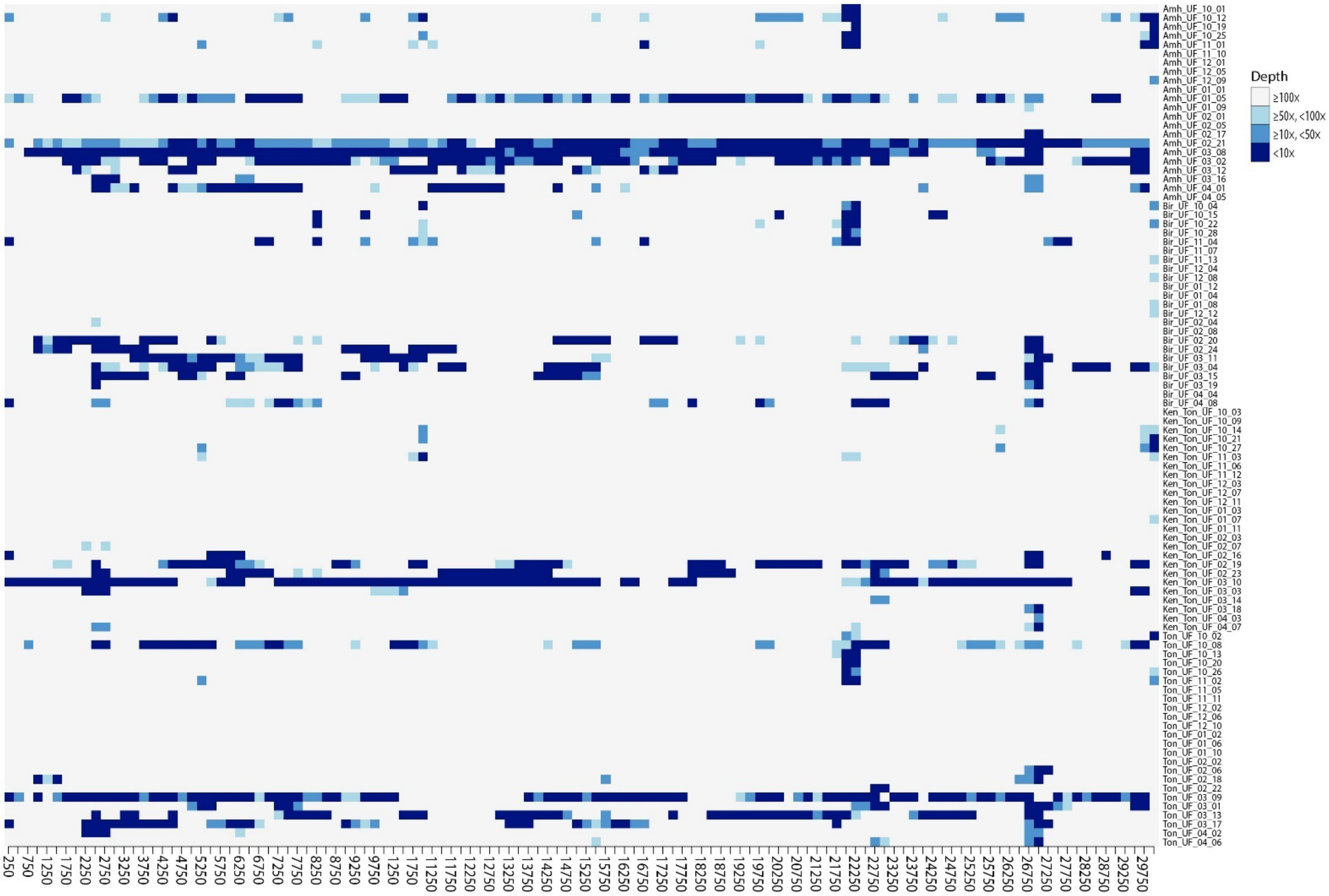


**Figure S7.** Heat map of read depths of SARS-CoV-2 whole-genome sequencing from wastewater samples. The depth was calculated for every 250-nucelotide region across the SARS-CoV-2 genome. Each row in the plot represents one wastewater sample, which was named "sewershed_SeqID". The Amherst, Bird Island, Kenmore-Tonawanda, and Tonawanda sewersheds are abbreviated as Amh, Bir, Ken_Ton, and Ton, respectively. The SeqID and sample details are provided in **Table S4**.

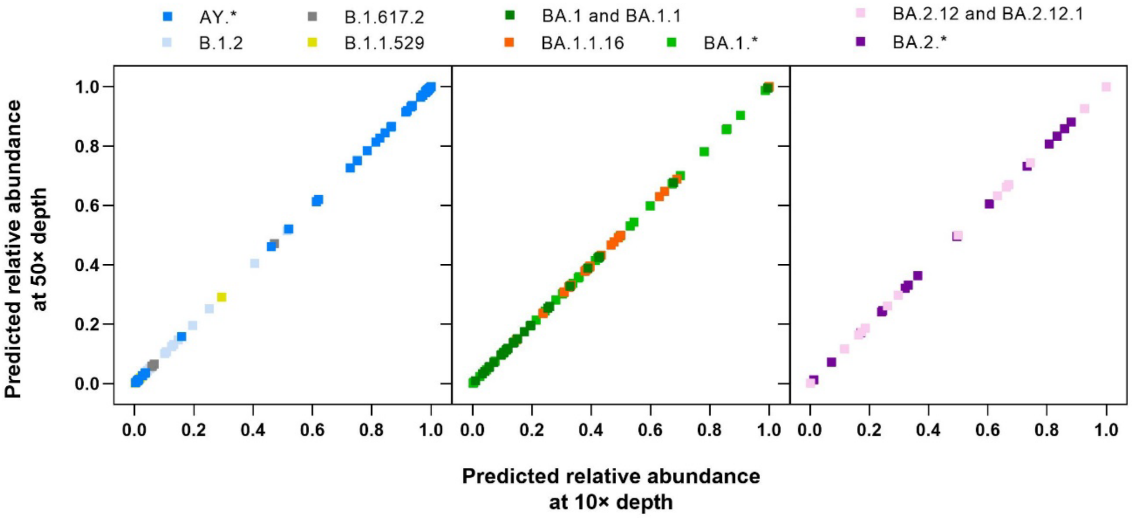


**Figure S8.** Comparison of relative abundances of SARS-CoV-2 lineages estimated by the sequencing data at depths of 10× and 50×. The Freyja pipeline[10] was used to estimate the relative abundances of different lineages.

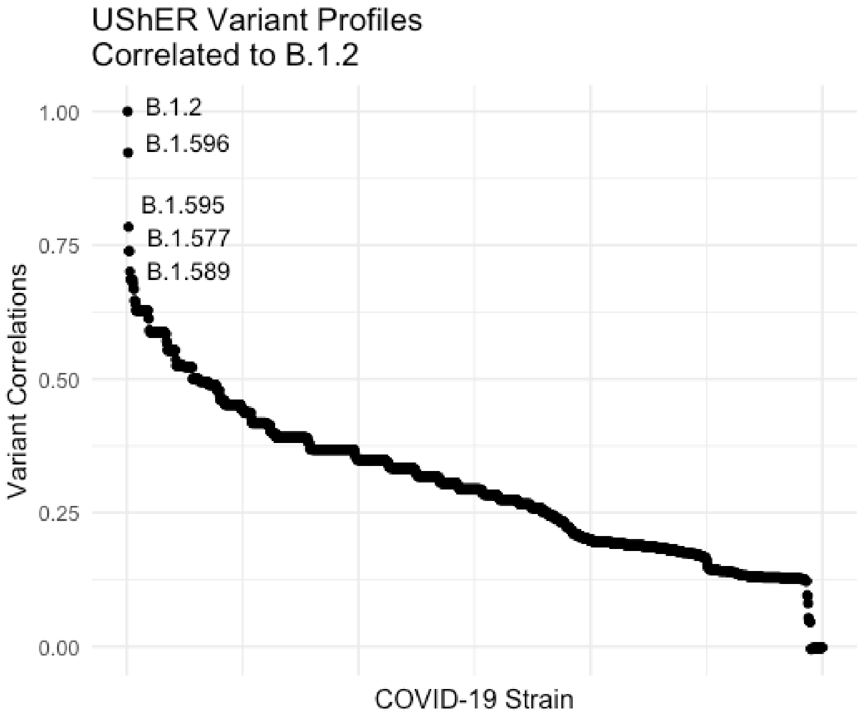


**Figure S9.** Pair-wise pearson correlation of B.1.2 lineage mutaton spectrum against all other lineage mutation spectra used in Freyja for calling lineages. The resulting pearson correlation values were sorted from highest to lowest. The lineages with the top five highest correlation values were labeled on the plot.

### REFERENCES


(1) Lu, X.; Wang, L.; Sakthivel, S. K.; Whitaker, B.; Murray, J.; Kamili, S.; Lynch, B.; Malapati, L.; Burke, S. A.; Harcourt, J.; et al. US CDC real-time reverse transcription PCR panel for detection of Severe Acute Respiratory Syndrome Coronavirus 2. *Emerg Infect Dis* **2020**, *26* (8), 1654. DOI: 10.3201/eid2608.201246.
(2) He, H.; Zhou, P.; Shimabuku, K. K.; Fang, X.; Li, S.; Lee, Y.; Dodd, M. C. Degradation and deactivation of bacterial antibiotic resistance genes during exposure to free chlorine, monochloramine, chlorine dioxide, ozone, ultraviolet light, and hydroxyl radical. *Environ Sci Technol* **2019**, *53* (4), 2013-2026. DOI: 10.1021/acs.est.8b04393.
(3) Lee, W. L.; Gu, X.; Armas, F.; Wu, F.; Chandra, F.; Chen, H.; Xiao, A.; Leifels, M.; Chua, F. J. D.; Kwok, G. W. C.; et al. Quantitative detection of SARS-CoV-2 Omicron BA.1 and BA.2 variants in wastewater through allele-specific RT-qPCR. *medRxiv* **2022**. DOI: 10.1101/2021.12.21.21268077.
(4) Lee, W. L.; Imakaev, M.; Armas, F.; McElroy, K. A.; Gu, X. Q.; Duvallet, C.; Chandra, F.; Chen, H. J.; Leifels, M.; Mendola, S.; et al. Quantitative SARS-CoV-2 Alpha variant B.1.1.7 tracking in wastewater by allele-specific RT-qPCR. *Environ Sci Technol Lett* **2021**, *8* (8), 675-682. DOI: 10.1021/acs.estlett.1c00375.
(5) Bustin, S. A.; Benes, V.; Garson, J. A.; Hellemans, J.; Huggett, J.; Kubista, M.; Mueller, R.; Nolan, T.; Pfaffl, M. W.; Shipley, G. L.; et al. The MIQE guidelines: minimum information for publication of quantitative real-time PCR experiments. *Clin Chem* **2009**, *55* (4), 611-622. DOI: 10.1373/clinchem.2008.112797.
(6) Bar-Or, I.; Weil, M.; Indenbaum, V.; Bucris, E.; Bar-Ilan, D.; Elul, M.; Levi, N.; Aguvaev, I.; Cohen, Z.; Shirazi, R.; et al. Detection of SARS-CoV-2 variants by genomic analysis of wastewater samples in Israel. *Sci Total Environ* **2021**, *789*, 148002. DOI: 10.1016/j.scitotenv.2021.148002.
(7) Izquierdo-Lara, R.; Elsinga, G.; Heijnen, L.; Munnink, B. B. O.; Schapendonk, C. M. E.; Nieuwenhuijse, D.; Kon, M.; Lu, L.; Aarestrup, F. M.; Lycett, S.; et al. Monitoring SARS-CoV-2 circulation and diversity through community wastewater sequencing, the Netherlands and Belgium. *Emerg Infect Dis* **2021**, *27* (5), 1405-1415. DOI: 10.3201/eid2705.204410.
(8) Perez-Cataluna, A.; Chiner-Oms, A.; Cuevas-Ferrando, E.; Diaz-Reolid, A.; Falco, I.; Randazzo, W.; Giron-Guzman, I.; Allende, A.; Bracho, M. A.; Comas, I.; et al. Spatial and temporal distribution of SARS-CoV-2 diversity circulating in wastewater. *Water Res* **2022**, *211*, 118007. DOI: 10.1016/j.watres.2021.118007.
(9) Fontenele, R. S.; Kraberger, S.; Hadfield, J.; Driver, E. M.; Bowes, D.; Holland, L. A.; Faleye, T. O. C.; Adhikari, S.; Kumar, R.; Inchausti, R.; et al. High-throughput sequencing of SARS-CoV-2 in wastewater provides insights into circulating variants. *Water Res* **2021**, *205*, 117710. DOI: 10.1016/j.watres.2021.117710.
(10) Karthikeyan, S.; Levy, J. I.; De Hoff, P.; Humphrey, G.; Birmingham, A.; Jepsen, K.; Farmer, S.; Tubb, H. M.; Valles, T.; Tribelhorn, C. E.; et al. Wastewater sequencing reveals early cryptic SARS-CoV-2 variant transmission. *Nature* **2022**, *609*, 101-108. DOI: 10.1038/s41586-022-05049-6.
